# Determinants of early antibody responses to COVID-19 mRNA vaccines in exposed and naive healthcare workers

**DOI:** 10.1101/2021.09.08.21263232

**Authors:** Gemma Moncunill, Ruth Aguilar, Marta Ribes, Natalia Ortega, Rocío Rubio, Gemma Salmeron, María José Molina, Marta Vidal, Diana Barrios, Robert A. Mitchell, Alfons Jimenez, Cristina Castellana, Pablo Hernández-Luis, Pau Rodó, Susana Méndez, Anna Llupià, Laura Puyol, Natalia Rodrigo Melero, Carlo Carolis, Alfredo Mayor, Luis Izquierdo, Pilar Varela, Antoni Trilla, Anna Vilella, Sonia Barroso, Ana Angulo, Pablo Engel, Marta Tortajada, Alberto L. García-Basteiro, Carlota Dobaño

## Abstract

**Background:** Two doses of mRNA vaccination have shown >94% efficacy at preventing COVID-19 mostly in naive adults, but it is not clear if the second dose is needed to maximize effectiveness in those previously exposed to SARS-CoV-2 and what other factors affect responsiveness.

**Methods:** We measured IgA, IgG and IgM levels against SARS-CoV-2 spike (S) and nucleocapsid (N) antigens from the wild-type and S from the Alpha, Beta and Gamma variants of concern, after BNT162b2 (Pfizer/BioNTech) or mRNA-1273 (Moderna) vaccination in a cohort of health care workers (N=578). Neutralizing capacity and antibody avidity were evaluated. Data were analyzed in relation to COVID-19 history, comorbidities, vaccine doses, brand and adverse events.

**Findings:** Vaccination induced robust IgA and IgG levels against all S antigens. Neutralization capacity and S IgA and IgG levels were higher in mRNA-1273 vaccinees, previously SARS-CoV-2 exposed, particularly if symptomatic, and in those experiencing systemic adverse effects. A second dose in pre-exposed did not increase antibody levels. Smoking and comorbidities were associated with lower neutralization and antibody levels. Among fully vaccinated, 6.3% breakthroughs were detected up to 189 days post-vaccination. Among pre-exposed non-vaccinated, 90% were IgG seropositive more than 300 days post-infection.

**Interpretation:** Our data support administering a single-dose in pre-exposed healthy individuals. However, heterogeneity of responses suggests that personalized recommendations may be necessary depending on COVID-19 history and life-style. Higher mRNA-1273 immunogenicity would be beneficial for those expected to respond worse to vaccination. Persistence of antibody levels in pre-exposed unvaccinated indicates maintenance of immunity up to one year.

**Funding:** This work was supported by Institut de Salut Global de Barcelona (ISGlobal) internal funds, in-kind contributions from Hospital Clínic de Barcelona, the Fundació Privada Daniel Bravo Andreu, and European Institute of Innovation and Technology (EIT) Health (grant number 20877), supported by the European Institute of Innovation and Technology, a body of the European Union receiving support from the H2020 Research and Innovation Programme. We acknowledge support from the Spanish Ministry of Science and Innovation and State Research Agency through the “Centro de Excelencia Severo Ochoa 2019-2023” Program (CEX2018-000806-S), and support from the Generalitat de Catalunya through the CERCA Program. L. I. work was supported by PID2019-110810RB-I00 grant from the Spanish Ministry of Science & Innovation. Development of SARS-CoV-2 reagents was partially supported by the National Institute of Allergy and Infectious Diseases Centers of Excellence for Influenza Research and Surveillance (contract number HHSN272201400008C). The funders had no role in study design, data collection and analysis, the decision to publish, or the preparation of the manuscript.

## INTRODUCTION

The unprecedented fast development of highly efficacious COVID-19 vaccines has changed the fate of the SARS-CoV-2 pandemic [1]. The COVID-19 vaccines from Pfizer/BioNTech (BNT162b2) and Moderna (mRNA-1273) manufacturers based on mRNA encoding the SARS-CoV-2 full-length spike (S) protein have shown vaccine efficacies of 95% and 94%, respectively, against COVID-19 disease after two doses in phase 3 trials [2, 3]. Both vaccines induce good immunogenicity [4–11] and excellent effectiveness in real world population after two doses [12–15] but lower effectiveness against variants of concern (VoC) after one dose [12,14,15] and against the Delta variant following two doses [16].

Unfortunately, vaccine production is limited, which has resulted in changes in immunization policies in many high- and medium-income countries, such as delays in the 2^nd^ dose, prioritization of naïve individuals over previously SARS-CoV-2 diagnosed individuals, or a single-dose for the latter ones. Nevertheless, evidence shows that previously infected individuals benefit from vaccination [4–11] and, therefore, the recommendation is to vaccinate the total population regardless of COVID-19 history [17, 18]. However, an increasing number of studies suggest that only one dose would be sufficient to mount an optimal antibody response in previously infected individuals, as a booster response is elicited [4–11]. This has led to the recommendation in some countries to provide only one dose to those previously diagnosed [19], and to the suggestion that two doses do not contribute to an additional improvement [7,8,11] or could even have a detrimental effect on the acquired immune response [7, 20]. This would also allow an increase to the global supply of doses available to low-income countries that suffer from vaccine shortages [21]. However, further evidence is needed to guide informed decisions as most of the studies include a small sample size and it is not clear whether it applies to all individuals [19, 22]. In Spain, a single-dose vaccination after at least 6 months post-infection is recommended for previously COVID-19 diagnosed individuals less than 65 years old [23] and has recently changed to 2 months in Catalonia [24]. In countries such as France or Germany, a single dose is also recommended for previously diagnosed healthy individuals [19]. However, other countries are still administering two doses to everyone and are even considering or starting to administer a 3^rd^ booster dose in light of declining antibody responses and the spread of highly contagious VoCs such as Delta, with potential immune escape and diminished vaccine effectiveness [16, 25].

The emergence of several fast-spreading variants since the end of 2020 may affect vaccination campaigns. Concern has been raised about the potential of some of the variants, which harbor mutations in S, to escape from neutralizing antibody immunity. Some studies have shown that antibodies from convalescent and vaccinated individuals are effective against the Alpha (B1.1.7) variant first identified in UK [26–29]. In contrast, Beta (B.1.351) and Gamma (P.1), first identified in South Africa and Brazil, respectively, have decreased sensitivity to neutralizing antibodies elicited by vaccination [27,28,30–33], but previous exposure induces higher cross-reactivity to variants [4, 9]. More recently, the Delta variant, which also presents mutations in S, was identified in India and quickly spread over the world, and early data show that it may have an even lower sensitivity to convalescent and vaccine induced antibodies [34, 35].

Since March 2020, we have followed up a cohort of 578 health care workers (HCW) at Hospital Clínic de Barcelona (HCB), Spain [36]. After 6 months of follow-up (October 2020) the cumulative prevalence of SARS-CoV-2 infection based on real-time reverse-transcriptase polymerase chain reaction (rRT-PCR) or serology data was 19.6%, but most of the infections occurred during the first wave of the pandemic [37]. Most of the infected individuals maintained IgG levels against S antigens and their neutralizing capacity up to 7 months [37]. In the present study, we evaluated the IgA, IgG and IgM levels and their neutralizing capacity early after vaccination with one or two doses of the BNT162b2 and mRNA-1273 vaccines, investigated the impact of previous SARS-CoV-2 infection history and antibody responses, and other variables like vaccine reactogenicity, comorbidities, or smoking habit, and report the breakthrough infections among fully vaccinated participants. In addition, we present the antibody kinetics to S and N antigens for up to 1-year post-infection for those individuals who have not been vaccinated.

## METHODS

### Study design, population and setting

The baseline study population included 578 randomly selected HCW from HCB who delivered care and services directly or indirectly to patients. Detailed characteristics of the study cohort have been described elsewhere [36–38]. Participants were recruited at the peak of the first wave of the pandemic in Spain (study month 0, M0) [36] and performed 2 additional visits at study month 1 (M1) and month 6 (M6) [37]. Participants with any previous evidence of SARS-CoV-2 infection were invited to participate at study months 3 (M3) [38] and 9 (M9) follow-up visits. All participants were invited again for a month 12 (M12) visit. At the M9 visit, 64 participants had already received one dose of the BNT162b2 (Comirnaty, Pfizer/BioNTech) or mRNA-1273 (Spykevax, Moderna) mRNA vaccines, with various times post vaccination. By M12, most of the participants had already received two doses of either vaccine and were invited to come for a cross-sectional visit 2 weeks (window 12-19 days) after the second dose was administered (N=342, BNT162b2 N=271, mRNA-1273 N =71). Finger prick blood was collected from the subset who visited at M9, and 10 mL of venous blood was collected from all participants at M12. Plasma was isolated and cryopreserved at - 80°C. We collected retrospective data on the vaccination dates, related adverse events (AEs) and COVID-19 infection and symptoms. We also collected information on new SARS-CoV-2 infection episodes in this cohort until 6 months after vaccination (M18) through the Occupational Health department at the HCB. We had previously collected demographic data and information on chronic medication and smoking habits. Data for each participant were collected and managed using REDCap version 8.8.2 hosted at ISGlobal through a standardized electronic questionnaire as previously described [36]. REDCap (Research Electronic Data Capture) is a secure, web-based software platform designed to support data capture for research studies [39, 40], providing 1) an intuitive interface for validated data capture; 2) audit trails for tracking data manipulation and export procedures; 3) automated export procedures for seamless data downloads to common statistical packages; and 4) procedures for data integration and interoperability with external sources.

### Quantification of antibodies to SARS-CoV-2

We measured IgA, IgG and IgM antibody levels (median fluorescence intensity, MFI) to different SARS-CoV-2 antigens using previously developed assays based on the quantitative suspension array technology Luminex (Supplementary Information) [37, 41]. The panel of antigens included the spike full length protein (S) (aa 1-1213 expressed in Expi293 and His tag-purified) produced at Center for Genomic Regulation (CRG, Barcelona), and its subregion S2 (purchased from SinoBiological, cat. No. 40590-V08B), the receptor-binding domain (RBD) kindly donated by the Krammer lab (Mount Sinai, New York) [42], the nucleocapsid (N) full length protein and the specific C-terminal region (both expressed in-house in *E. coli* and His tag-purified) [43], and the full length S proteins of 3 VoCs (purchased from Acro Biosystems): Alpha (B.1.1.7; cat. No. SPN-C52H6), Beta (B.1.351; cat. No. SPN-C52Hk) and Gamma (P.1; cat. No. PN-C52Hg). Plasma samples were tested at 1:500 dilution for the 3 isotypes, and additionally at 1:5000 for IgG to avoid saturated levels in the vaccinated participants.

### Neutralizing antibodies

For feasibility reasons, we selected 165 samples from the study visit M12 with a balanced representation of BNT162b2 and mRNA-1273 vaccinees and non-vaccinated participants (previously exposed and naive individuals). We already had pre-vaccination neutralization data from 33 of the selected 165 individuals [37]. Plasma neutralizing capacity was assessed as the percentage of inhibition of RBD binding to ACE2 receptor and was measured through a flow cytometric-based assay that correlates with a validated pseudovirus neutralization assay [37]. Briefly, a murine stable cell line expressing the ACE2 receptor was incubated with RBD-mFc fusion protein, composed of RBD fused to the Fc region of murine IgG1, previously exposed to the different plasma samples at a 1:400 dilution. Cells were stained with anti-mouse IgG-PE, washed, and analyzed by flow cytometry using standard procedures. Study samples were tested alongside 30 negative pre-pandemic controls, in duplicates.

### Antibody avidity

For feasibility, a subset of 58 M12 samples from BNT162b2 and mRNA-1273 vaccinated participants (48 naive and 10 exposed), were randomly selected for the avidity assay. Antibody avidity was determined as the percentage of IgA and IgG levels against RBD, S and S2 antigens measured incubating samples with a chaotropic agent (urea 4M, 30 min at room temperature) over the IgA and IgG levels measured in the same samples without chaotropic agent. Antibody levels with and without chaotropic agents were measured in plasma samples (dilution 1:5000) using the Luminex assay described above.

### Statistical analysis

MFIs were log_10_-transformed. In vaccinated participants MFIs for S-related antigen IgGs correspond to the 1:5000 dilution, except in plots where we compare pre and post vaccination levels, in which the 1:500 dilution was used. Any other MFIs correspond to the dilution 1:500, and for seropositivity calculations, only 1:500 dilution was used.

Assay positivity cutoffs specific for each isotype and analyte were calculated as 10 to the mean plus 3 standard deviations (SD) of log_10_-transformed MFI of 128 pre-pandemic controls. Results were defined as undetermined when the MFI levels for a given isotype-analyte were between the positivity threshold and an upper limit defined as 10 to the mean plus 4.5 SD of the log_10_-transformed MFIs of 128 pre-pandemic samples, and no other isotype-antigen combination was above the positivity cutoff, and the participant did not have any previous evidence of seropositivity or rRT-PCR positivity.

Analysis of antibody levels after the second dose included only data from samples collected 12-19 days after vaccination, while for the first dose we included data from all samples collected 7 or more days after vaccination, as no previous visit window was established. Groups were compared using the Wilcoxon Sum Rank test for continuous non-parametric variables and with the Wilcoxon Signed-Sum Rank Test for paired continuous data.

Correlations between continuous variables were analyzed using linear regression models and Spearman’s rank test. Locally estimated scatterplot smoothing (LOESS) plots were used to visualize trends in antibody levels over days post vaccination, days post-symptom onset (PSO) or post rRT-PCR diagnosis.

Univariable and multivariable linear regression models were fitted to assess factors associated with antibody responses to SARS-CoV-2 RBD and S antigens and their neutralization capacity (%) after vaccination among exposed and naive individuals, and overall. Models having both exposed and naive participants included the following independent variables: sex, age, days since first dose administration, days since second dose administration, smoking habits, chronic medication, presence of baseline illness (heart and liver disease, diabetes, chronic respiratory and renal disease, cancers and autoimmune and other immunological disorders), antibody levels (log_10_ MFI) to endemic common cold human coronaviruses (HCoV: 229E, NL63, OC43, and HKU1) at M6, vaccine type, and presence of systemic or local AEs (systemic AEs included fever, arthralgia, fatigue, chills, muscle pain and headache, while local AEs included pain, erythema and/or swelling at the injection site or swollen glands near the injection site) after 1^st^ or 2^nd^ vaccine dose. In addition, the predictor variable “presence of any COVID-19 symptom (fatigue, cough, dyspnea and other respiratory symptoms, anosmia or ageusia, sore throat, fever, rhinorrhea, headache, chills and digestive symptoms)” was included in models having only exposed participants. Predictor variables that had a P-value of 0.2 in the univariable models were selected for stepwise multivariable models performed with the function stepAIC (R package MASS). The betas obtained in each model for each of the predictor variables were transformed into a percentage of antibody increase for easier interpretation. For continuous log_10_-transformed variables (log-log model) the beta transformed value (%) was calculated with the formula ((10^(beta*log_10_(1.1)))-1)*100. This represents the effect (in percentage) on IgG levels of a 10% increase in the corresponding predictor variable. For categorical predictor variables (log-linear models), the beta transformed value (%) was calculated with the formula ((10^beta)-1)*100. This gives the difference (in percentage) in IgG levels between the reference and the study group. A P-value of ≤ 0.05 was considered statistically significant and 95% confidence intervals (CI) were calculated for all estimates. We performed the statistical analysis in R version 4.0.3 (packages tidyverse, ggpubr and MASS) [44–46].

### Ethics

Written informed consent was obtained from all study participants prior to study initiation. The study was approved by the Ethics Committee at HCB (references HCB/2020/0336 and HCB/2021/0196).

## RESULTS

### Characteristics of study participants

From the 578 participants recruited at baseline, 446 came to visits at M9 and/or M12, with 414 participants sampled at M12. We measured the levels of IgA, IgG and IgM to SARS-CoV-2 antigens in blood samples from both visits. Of the 414 HCW visited at M12, 360 (81%) had received one (N= 18, 1 BNT162b2 and 17 mRNA-1273) or two doses (N= 342) of the mRNA vaccines. Seventy-six percent of the 360 HCW received BNT162b2 and 24% mRNA-1273 (Table 1). Most of the study participants were females (73%) and had a mean age of 42.7 (SD 11.65) years. Around 20% had underlying comorbidities and 22% were under chronic medication (Table 1). Thirty-two per cent of all participants and 22% of those vaccinated had previously been infected by SARS-CoV-2 according to rRT-PCR or serology data (Table 1). Seventy-three per cent of the participants had AEs to vaccination, systemic in 28% after one dose and 68% after two doses (Table 1). Among the 159 participants fully vaccinated with two doses, 10 (6.3%) vaccine breakthroughs were detected by rRT-PCR after 15 days post-second dose with a median of 144.5 days (49-189 days) post-vaccination. Among the 53 individuals non-vaccinated at M12, 4 (7.5%) had a SARS-CoV-2 infection in the same period of time.

**Table 1.** Characteristics of study participants.

| Variable | N at M9<br>and/or M12 | % or mean<br>(SD) | N Vaccinated<br>at M12 | % or mean<br>(SD) |
| --- | --- | --- | --- | --- |
| **Sex** | 446<sup>a</sup> |  | 360 |  |
| Male | 119 | 26.70% | 94 | 26.10% |
| Female | 327 | 73.30% | 266 | 73.90% |
| **Age (years)** | 446 | 42.7 (11.65) | 360 | 43.2 (11.77) |
| **Job function** | 446 |  | 360 |  |
| Nurses and auxiliary health<br>professionals | 229 | 51.30% | 177 | 49.20% |
| Physicians and psychologists | 105 | 23.50% | 89 | 24.70% |
| Laboratory and other technicians | 34 | 7.60% | 26 | 7.20% |
| Other<sup>b</sup> | 78 | 17.50% | 68 | 18.90% |
| **Comorbidities<sup>c</sup>** | 446 |  | 360 |  |
| No | 356 | 79.80% | 289 | 80.30% |
| Yes | 90 | 20.20% | 71 | 19.70% |
| **Chronic medication** | 446 |  | 360 |  |
| No | 350 | 78.5% | 279 | 77.5% |
| Yes | 96 | 21.5% | 81 | 22.5% |
| **Smoker** | 445 |  | 360 |  |
| No | 346 | 77.80% | 280 | 77.80% |
| Yes | 99 | 22.20% | 80 | 22.20% |
| **Number people in the**<br>**household** | 446 | 2.74 (1.173) | 360 | 2.76 (1.184) |
| **Involved in clinical care** | 446 |  | 360 |  |
| No | 97 | 21.70% | 88 | 24.40% |
| Yes | 349 | 78.30% | 272 | 75.60% |
| **Number children co-living** | 445 | 0.45 (0.8) | 360 | 0.45 (0.8) |
| **Vaccine type** |  |  | 360 |  |
| BNT162b2 (Pfizer/BioNTech) |  |  | 272 (n= 1<br>one dose) | 75.60% |
| mRNA-1273 (Moderna) |  |  | 88 (n= 17<br>one dose) | 24.40% |
| **Number doses received** |  |  | 360 |  |
| 1 |  |  | 18 | 5% |
| 2 |  |  | 342 | 95% |
| **Previously exposed** | 446 |  | 360 |  |
| No | 303 | 67.90% | 282 | 78.30% |
| Yes | 143 (92<br>symptomatic) | 32.10%<br>(64.33%<br>symptomatic) | 78 (43<br>symptomatic) | 21.70%<br>(55.13%<br>symptomatic) |
| **Adverse events after 1<sup>st</sup> dose** |  |  | 358 |  |
| Local/No Adverse Events |  |  | 257 | 71.79% |
| Systemic<sup>d</sup> |  |  | 101 | 28.21% |
| **Adverse events after 2<sup>nd</sup> dose** |  |  | 346 |  |
| Local/No Adverse Events |  |  | 110 | 31.79% |
| Systemic<sup>c</sup> |  |  | 236 | 68.21% |

<sup>a</sup> N at only M12 was 414
<sup>b</sup> Other include administration, accounting, information technology, cleaning, kitchen and maintenance staff.
<sup>c</sup> Comorbidities include: heart and liver disease, diabetes, chronic respiratory and renal disease, cancers and autoimmune and other immunological disorders.
<sup>d</sup> Systemic adverse events include fever, arthralgia, fatigue, chills, muscle pain and headache, while local adverse events include pain, erythema and/or swelling at the injection site or swollen glands near the injection site.
M: Study month; SD: Standard deviation.

### Vaccination elicits high but variable antibody levels against SARS-CoV-2 and VoC

After 7 to 72 days post one dose of the BNT162b2 or the mRNA-1273 vaccines, 92.2% (59/64) participants were seropositive, and seropositivity increased to 95.9% (47/49) when excluding samples from less than 10 days post-vaccination. IgA and IgG levels against all S antigens tested (RBD, S full length and S2) increased in most of the participants after one dose, albeit at very heterogeneous levels (Fig.1a). After 2 doses of the BNT162b2 or the mRNA-1273 vaccines (12-19 days post-vaccination), all participants were seropositive with the exception of a participant receiving the BNT162b2 who had renal insufficiency and was under corticoids and immunomodulatory cytokine treatments (Fig. 1b). IgA, IgG and IgM levels (Fig. 1b) and neutralization capacity (Fig. 2) increased in all individuals after the two doses compared to pre-vaccination but at varying levels.

**Figure 1.**
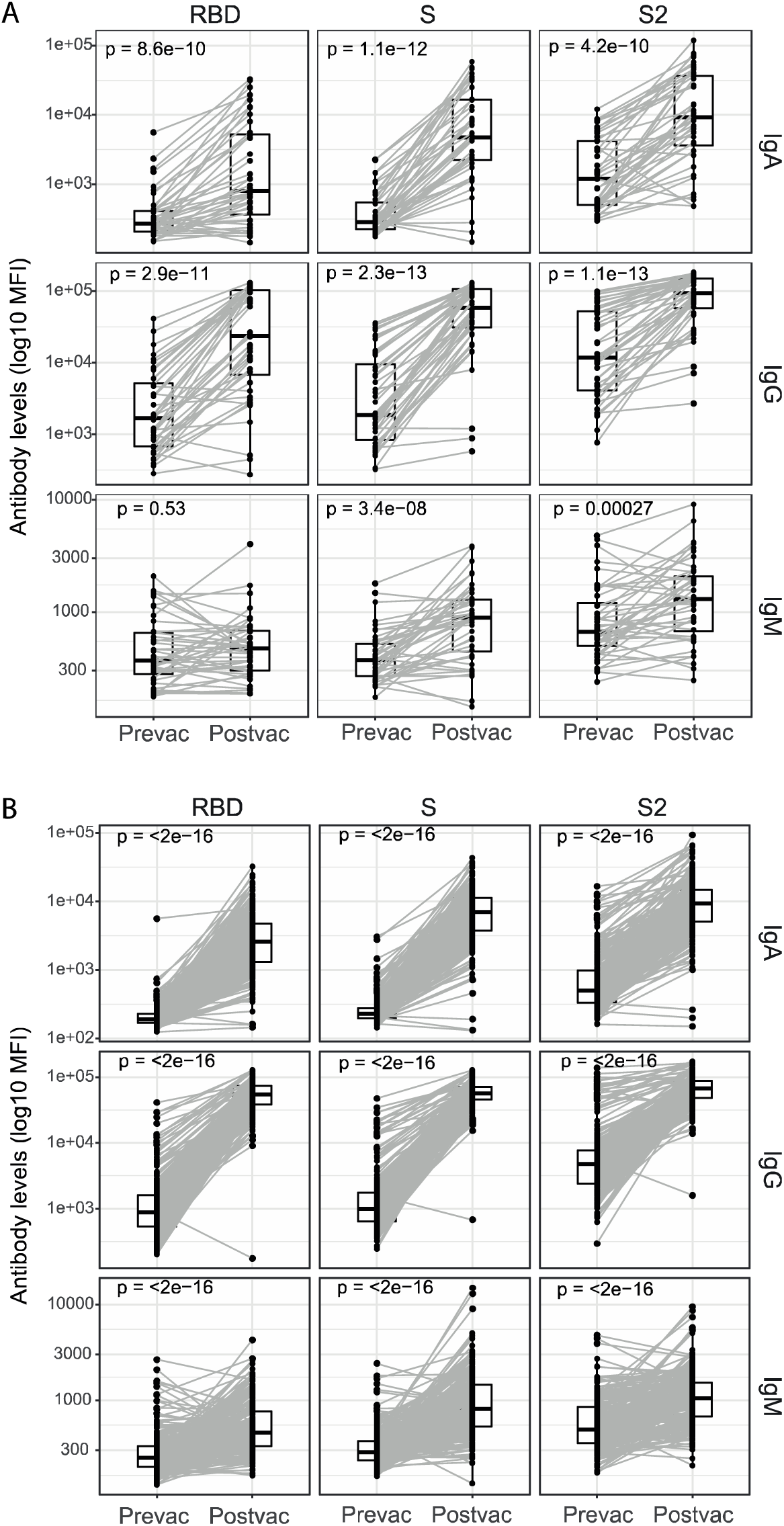
Pre- and post-vaccination antibody levels after 1 dose and 2 doses of the COVID-19 mRNA vaccines. Plots show IgA, IgG and IgM levels (median fluorescence intensity, MFI) against the receptor-binding domain (RBD) of the SARS-CoV-2 Spike glycoprotein (S), the S protein and its subunit S2 at pre- and post-vaccination after 1 dose (N=44) (A) and 2 doses (N=253) (B). All plasma samples were analyzed at 1:500 dilution. Pre-vaccination samples were collected at study month 6 for those who were already vaccinated at month 9, and at month 9 for those vaccinated at month 12. Post-vaccination samples analyzed are those collected >10 days after the 1^st^ dose (A) and 2 weeks after the 2^nd^ dose (B). Paired samples are joined by grey lines. The center line of boxes depicts the median of MFIs; the lower and upper hinges correspond to the first and third quartiles; the distance between the first and third quartiles corresponds to the interquartile range (IQR); whiskers extend from the hinge to the highest or lowest value within 1.5 × IQR of the respective hinge. Wilcoxon signed-rank test was used to assess statistically significant differences in antibody levels between pre- and post-vaccination.

**Figure 2.**
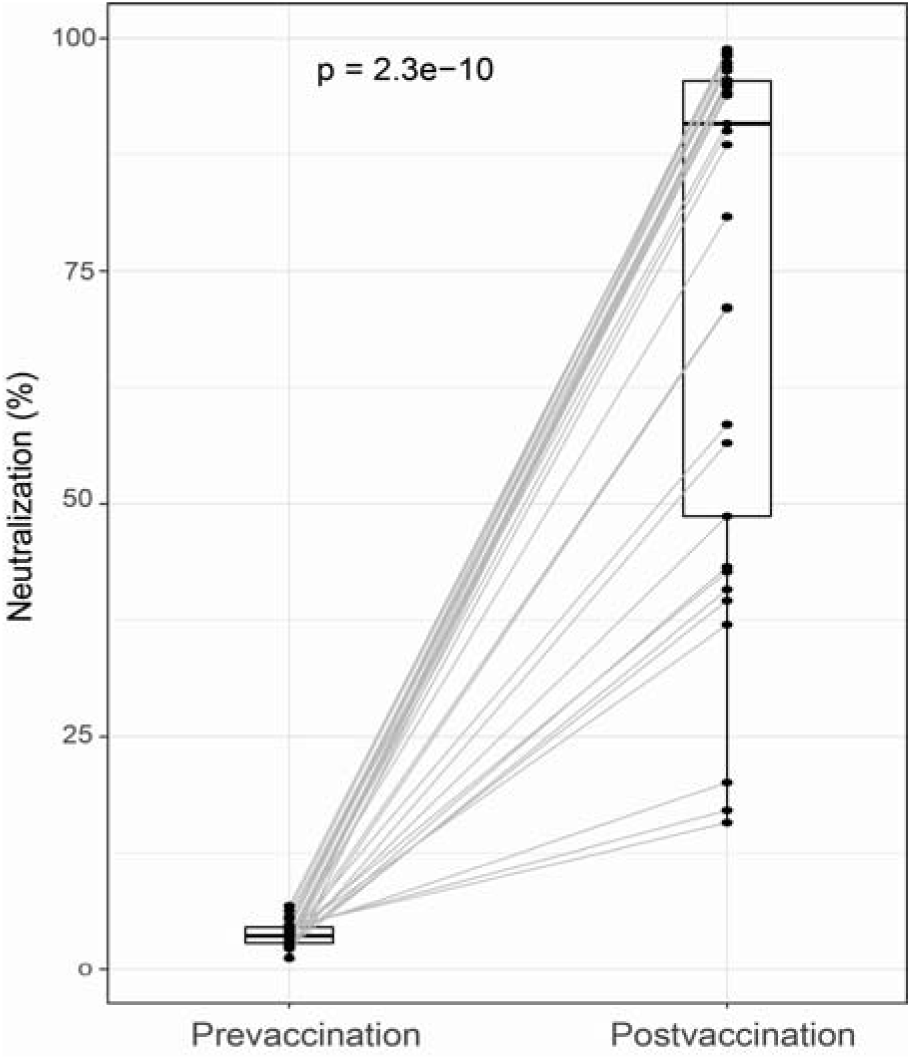
Neutralizing capacity of plasma samples before and after COVID-19 mRNA vaccination. Dots depict antibody neutralizing capacity, as a percentage of RBD-ACE2 binding inhibition by plasma samples from 33 participants. Paired samples are joined by grey lines. The center line of boxes depicts the median of the neutralization percentage; the lower and upper hinges correspond to the first and third quartiles; the distance between the first and third quartiles corresponds to the interquartile range (IQR); whiskers extend from the hinge to the highest or lowest value within 1.5 × IQR of the respective hinge. Wilcoxon signed-rank test was used to assess statistically significant differences between pre- and post-vaccination neutralization. Pre-vaccination levels correspond to visit M6. Pre- vaccination samples were analyzed at a 1:50 dilution while post vaccination were at 1:400. We standardized the post-vaccination results to make them comparable by dividing them by 8.

IgG antibodies produced after two doses in all seropositive individuals recognized the S full length from the Alpha, the Beta and the Gamma VoCs (Fig. S1). However, the odds of being IgM seronegative were 4.7 times higher for the Gamma variant, 3.8 times higher for the Beta variant and 2.5 times higher for the Alpha variant than for the wild-type.

### Association of previous SARS-CoV-2 exposure with antibodies post-vaccination

Previously SARS-CoV-2 infected individuals produced higher IgA, IgG and IgM levels against the S antigens RBD, S full length, and S2 after 1 (5.53 median fold-change increase for IgG, all antigens pooled) and 2 doses (1.36 median fold-change increase for IgG, all antigens pooled) of the vaccine than naive participants (Fig. 3). Kinetics after vaccination (Fig. S2) also show that vaccinated people who were previously exposed mounted higher antibody levels than naive individuals. Differences were larger after a 1^st^ dose than after 2 doses (Fig. 3-Fig. S2). Indeed, in previously infected individuals, antibody levels after the 2^nd^ dose were similar to levels after the 1^st^ dose with the exception of IgG against S2 that were lower after the 2^nd^ dose (Fig. 3), while for unexposed individuals, antibody levels clearly reached their maximum after the second dose. Differences in antibody levels between pre-exposed and unexposed individuals were similar for the S proteins of the Alpha, Beta and Gamma VoCs (Fig. S2).

**Figure 3.**
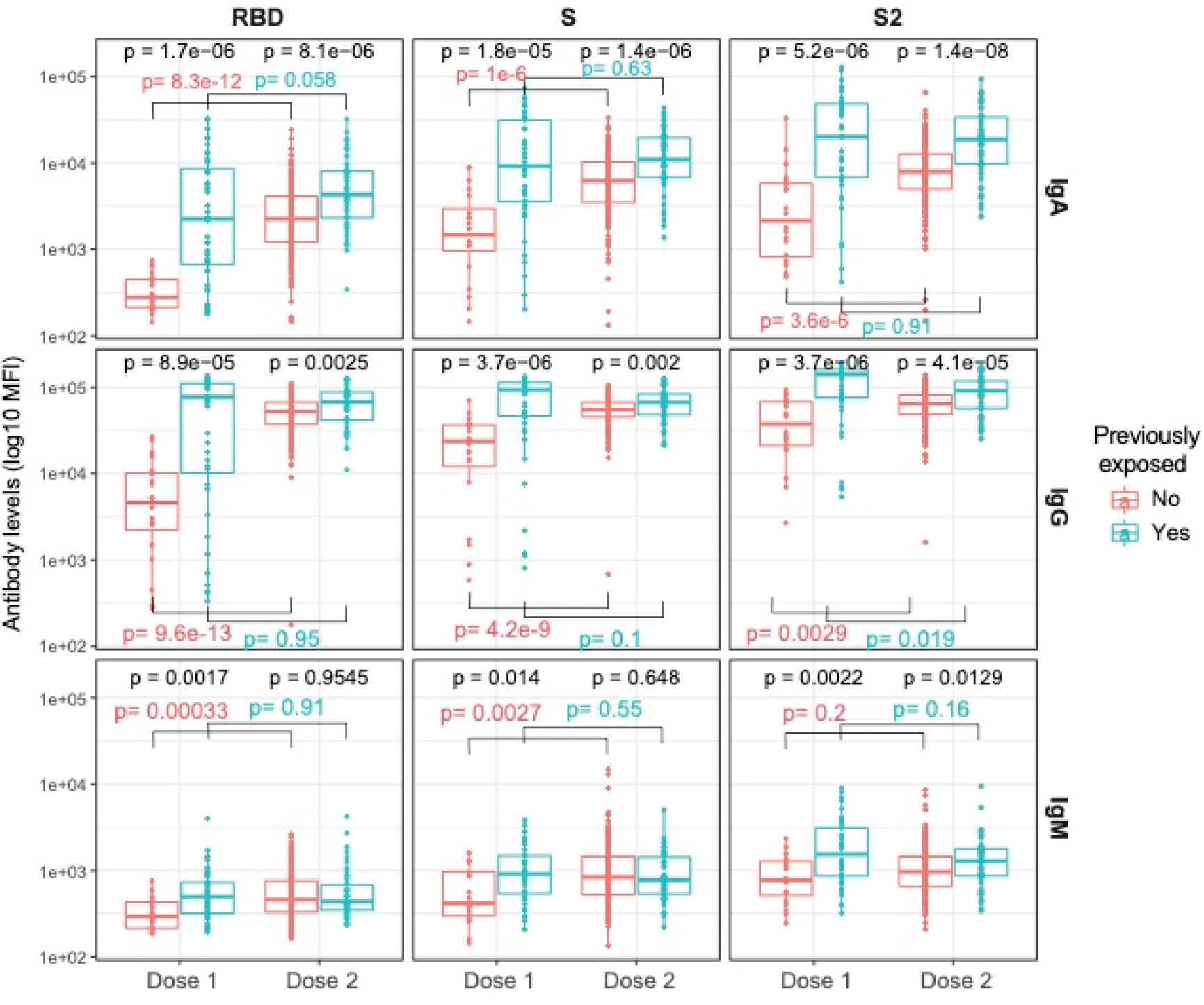
Antibody levels against S antigens after one and two doses of mRNA vaccines in previously SARS-CoV-2 infected and uninfected individuals. Plots show IgA, IgG and IgM levels (log_10_ MFI) against the receptor-binding domain (RBD) of the SARS-CoV-2 Spike glycoprotein (S), the S protein and its subunit S2 after 1 dose (N=64, 20 naive and 44 pre-exposed) and 2 doses (N=263, 211 naive and 52 pre-exposed). Post-vaccination samples analyzed were those collected >7 days after the 1^st^ dose and 2 weeks after the 2^nd^ dose. The center line of boxes depicts the median of MFIs; the lower and upper hinges correspond to the first and third quartiles; the distance between the first and third quartiles corresponds to the interquartile range (IQR); whiskers extend from the hinge to the highest or lowest value within 1.5 × IQR of the respective hinge. Wilcoxon rank test was used to assess statistically significant differences in antibody levels between naive and pre-exposed participants for a same dosage, and between 1^st^ and 2^nd^ dose into each group. We selected all dilutions at 1:500 to make levels comparable.

Antibody neutralization capacity after 2 vaccine doses was higher in pre-exposed than naive individuals (Fig. 4a). Similarly, the avidity of IgA and IgG in pre-exposed individuals after the 2^nd^ dose was higher compared to unexposed individuals (Fig. 4b). The plasma neutralization capacity positively and strongly correlated with IgG levels (for IgG RBD and S: rho=0.81-0.76 in naive and 0.83-0.84 in pre-exposed, p<0.001; Fig. S3) and moderately with IgA levels, particularly for RBD and in previously exposed participants (for IgA RBD: rho=0.45 p=0.002 in naïve and rho=0.61 in pre-exposed p<0.001; Fig. S3).

**Figure 4.**
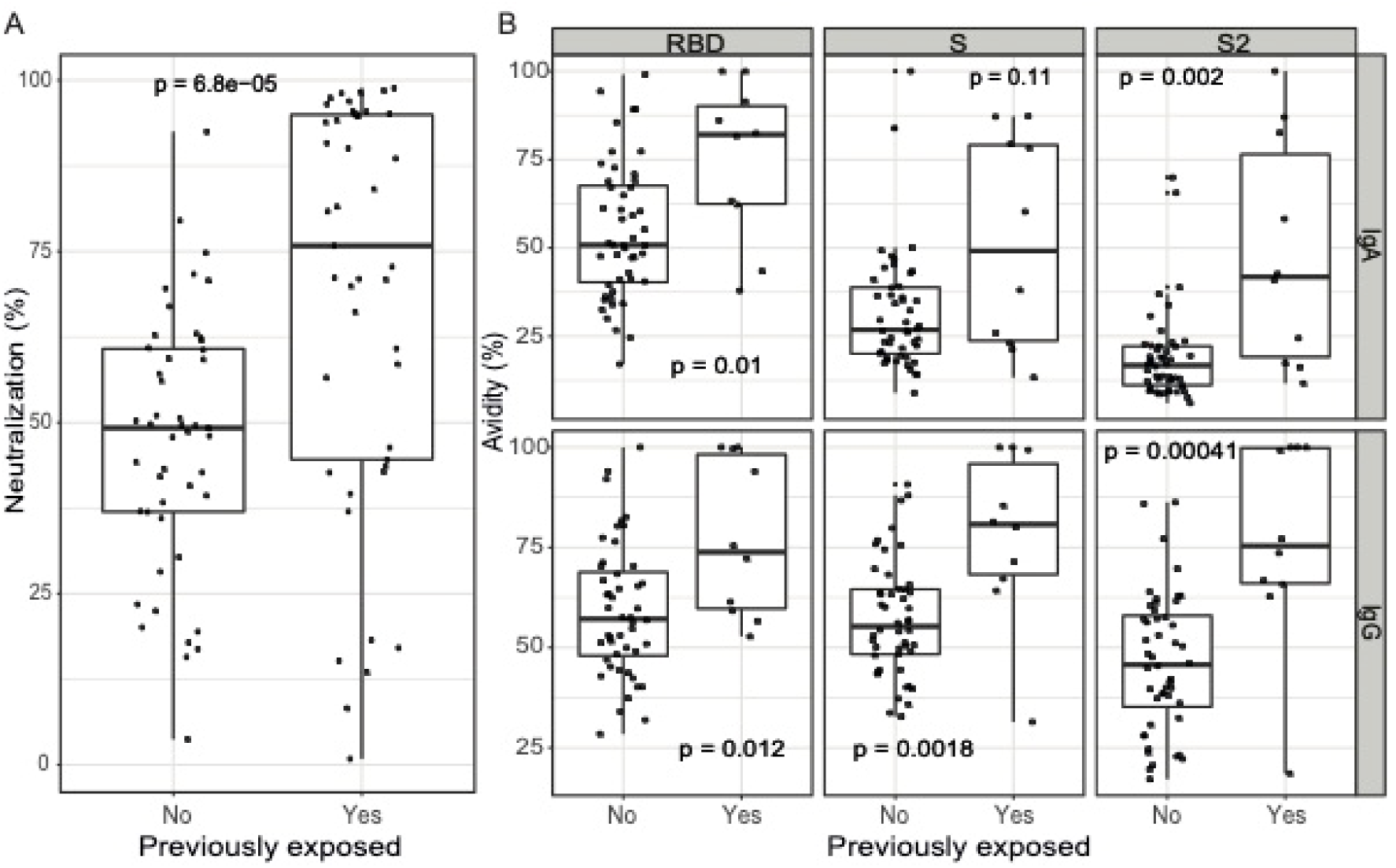
Antibody neutralization capacity and avidity after two doses of COVID-19 mRNA vaccines in naive and pre-exposed participants. A) Antibody neutralizing capacity, as a percentage of RBD-ACE2 binding inhibition by plasma samples assayed at 1:400 dilution (N=92, 47 naive and 45 pre-exposed). B) Antibody avidity, as % of IgA and IgG levels against RBD, S and S2 antigens measured incubating samples with a chaotropic agent over the IgA and IgG levels measured in the same samples without chaotropic agent, all at 1:5000 dilution (N=58, 48 naive and 10 pre-exposed). The center line of boxes depicts the median of MFIs; the lower and upper hinges correspond to the first and third quartiles; the distance between the first and third quartiles corresponds to the interquartile range (IQR); whiskers extend from the hinge to the highest or lowest value within 1.5 × IQR of the respective hinge. Wilcoxon rank test was used to assess statistically significant differences in antibody neutralization and avidity between naive and pre-exposed participants.

### Prior SARS-CoV-2 infection and antibody levels affect vaccine responses

When comparing responses between HCW who had the infection more than 11 months vs less than 11 months before vaccination, the first ones induced higher levels of IgA and IgG (Fig. S4). However, when using different cutoff values for the time passed between infection and vaccination, there were no differences in antibody levels induced by the vaccines.

Previously exposed participants who had symptoms during infection produced higher IgA and IgG levels against RBD and S2 after 2 vaccine doses than asymptomatic participants (Fig. S5a). Symptomatic individuals also had higher IgA and IgG levels against S full length VoCs (Fig. S5b) and had higher plasma neutralization capacity (Fig. S5c). In contrast, an inverse tendency was observed for IgM (Fig. S5a,b).

Pre-vaccination IgG and IgA levels in exposed participants positively and moderately correlated with antibody levels post-1^st^ and 2^nd^ dose (Fig S6a-b). Pre-vaccination IgM levels also correlated with post-vaccination levels but to a lesser extent. Antibody levels elicited after one dose positively correlated with the antibody levels elicited after the 2^nd^ dose among previously exposed (Fig. S6c).

### mRNA-1273 vaccine elicits higher antibody responses than BNT162b2

Two doses of the mRNA-1273 vaccine elicited higher IgA and IgG levels against RBD, S full length and the S2 subunit, and of higher neutralizing capacity and avidity, than two doses of the BNT162b2 vaccine (Fig. 5). Similarly, IgA and IgG levels against the S full length protein of the tested VoCs were higher after mRNA-1273 than BNT162b2 vaccination (Fig.5).

**Figure 5.**
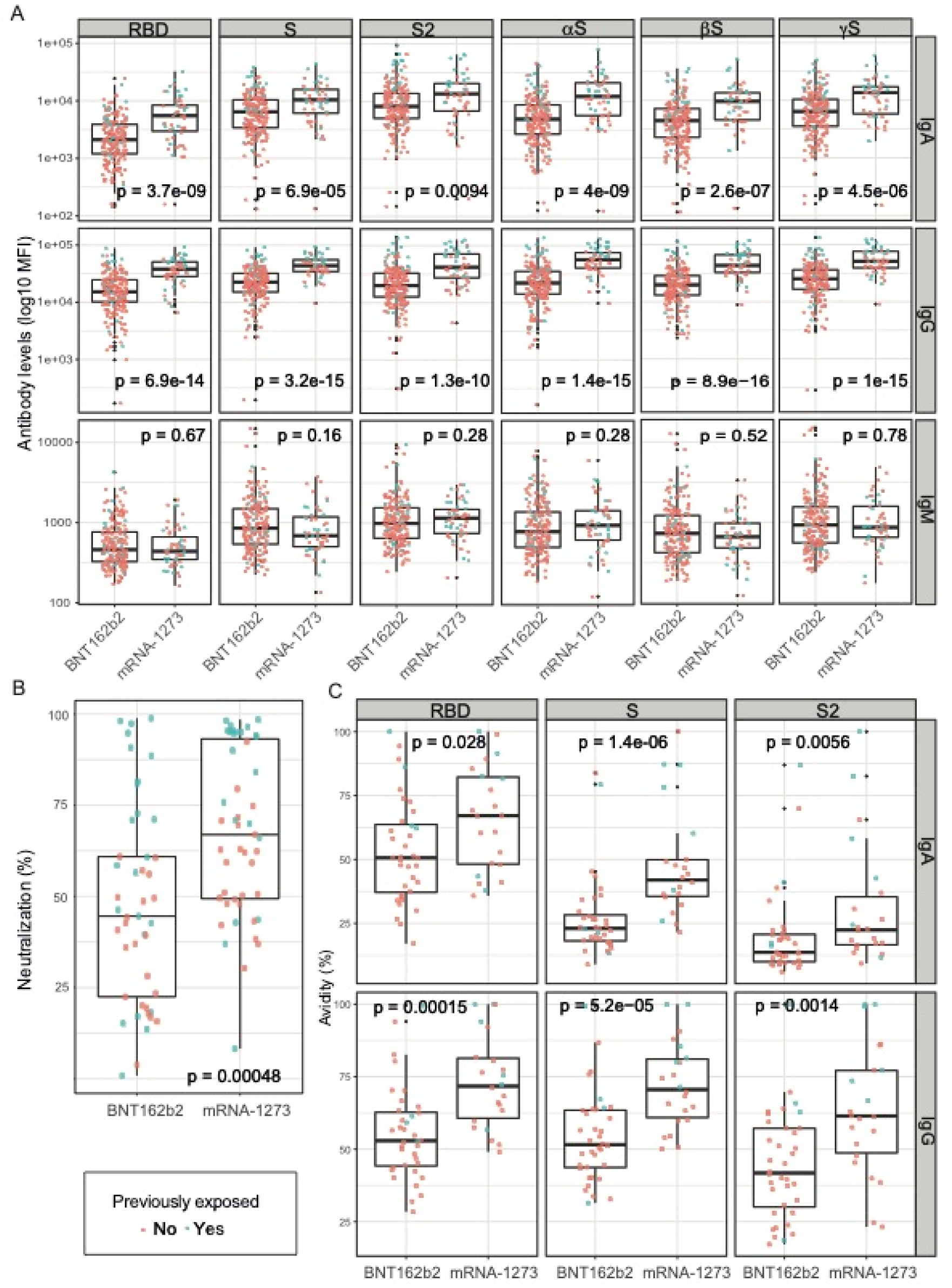
Comparison of antibody levels, neutralization and avidity between the two COVID-19 mRNA vaccines after two doses. A) Antibody levels elicited by BNT162b2 and mRNA-1273 among naive and pre-exposed participants (N = 263, 205 BNT162b2 / 56 mRNA-1273, 211 naïve, 52 exposed). Plasma samples were analyzed at 1:5000 dilution for IgG and 1:500 for IgA/IgM. B) Plasma neutralization capacity elicited by BNT162b2 and mRNA-1273 among naive and pre-exposed participants (N=92, 45 BNT162b2r/47 mRNA-1273, 47 naive, 45 exposed). Plasma dilution used was 1:400. C) Antibody avidity elicited by BNT162b2 vs mRNA-1273 among naive and pre-exposed participants (N=58, 36 BNT162b2 and 22 mRNA-1273, 48 naive, 10 pre-exposed). Plasma dilution used was 1:5000 for IgG and 1:500 for IgA. Red and green dots correspond to naive and pre-exposed participants, respectively.

### AEs after vaccination are associated with induction of higher antibody levels

Having had systemic AEs after 1^st^ dose was associated with higher levels of IgA and IgG against RBD and IgG against the S protein from the wild-type and the VoCs compared to having no or only local AEs (Fig. S7a). Similarly, having had systemic AEs after the 2^nd^ dose was associated with higher IgA, IgG and IgM levels to almost all S antigens than not having or only local AEs (Fig. S7b). Systemic AEs were also positively associated with higher neutralization capacity and avidity after the 2^nd^ dose (Fig. S7c-d).

### Factors independently associated with vaccine antibody responses after one dose

In univariable models, previously exposed HCW had 839% (260-2347, 95%CI; P-value<0.001) higher IgG S levels than naive HCW after a single-dose of the vaccines. BNT162b2 vaccination was associated with 78% lower IgG S levels (38-93, 95%CI; P-value=0.005) compared to mRNA-1273, whereas having had systemic AEs in contrast to local AEs or no AEs and days since 1^st^ dose were significantly and positively associated with 350% (56-1202, 95% CI; P-value=0.006) and 10% (2-19, 95% CI; P-value=0.13) higher IgG S levels, respectively.

In a stepwise multivariable model, these variables were retained but only previous exposure to SARS-CoV-2 and systemic AEs after vaccination were statistically significant (Table 2). In addition, smoking was associated with significantly less IgG S levels (63%, 6-85, 95% CI; P-value=0.038).

**Table 2.** Step-wise multivariable models assessing the impact of several variables on the IgG levels against the S full length protein induced after one (>7 days) and two doses of mRNA vaccines (12-19 days post-vaccine).

|  | Beta (%)<sup>a</sup> | Lower<br>95% CI | Upper<br>95% CI | P-value |
| --- | --- | --- | --- | --- |
| **One dose** |  |  |  |  |
| **naive + exposed participants N=55** |  |  |  |  |
| Smoker | -62.45 | -85.07 | -5.58 | **0.038** |
| SARS-CoV-2 pre-exposure | 526.74 | 135.96 | 1564.75 | **0.000** |
| BNT162b2 (ref: mRNA-1273) | -11.08 | -74.56 | 210.88 | 0.851 |
| Days since dose 1 | 5.79 | -2.48 | 14.77 | 0.171 |
| Systemic AEs dose 2 (ref: local/no AEs) | 171.88 | -2.05 | 654.64 | 0.055 |
| **Two doses** |  |  |  |  |
| **Naive + exposed participants N=262** |  |  |  |  |
| Sex (ref: male) | 19.33 | -0.31 | 42.85 | 0.054 |
| Comorbidities | -17.55 | -32.11 | 0.13 | 0.052 |
| SARS-CoV-2 pre-exposure | 38.18 | 12.97 | 69.02 | **0.002** |
| BNT162b2 (ref: mRNA-1273) | -43.27 | -53.58 | -30.66 | **<0.001** |
| Systemic AEs dose 2 (ref: local/no AEs) | 23.27 | 3.70 | 46.54 | **0.018** |
| **Naive only N=211** |  |  |  |  |
| Sex (ref: male) | 21.97 | -0.17 | 49.03 | 0.052 |
| Age continuous | -39.64 | -69.42 | 19.13 | 0.145 |
| BNT162b2 (ref: mRNA-1273) | -45.42 | -57.19 | -30.41 | **<0.001** |
| Systemic AEs dose 2 (ref: local/no AEs) | 28.61 | 5.87 | 56.24 | **0.012** |
| **Exposed only N=52** |  |  |  |  |
| Smoker | -35.40 | -56.83 | -3.34 | **0.034** |
| Comorbidities | -55.05 | -69.68 | -33.36 | **0.001** |
| BNT162b2 (ref: mRNA-1273) | -48.76 | -61.73 | -31.38 | **0.001** |
| Symptomatic (ref. Asymptomatic) | 38.72 | -1.28 | 94.95 | 0.059 |
| IgG levels against HKU1 N antigen | -1.31 | -2.91 | 0.31 | 0.11 |

<sup>a</sup> Beta transformed values to a percentage for an easier interpretation of variables effect

### Factors independently associated with vaccine antibody responses after two doses

In univariable models, we found that males had higher IgG levels against S full length protein than females, and that IgG levels decreased by age in unexposed vaccinated participants, but not in exposed participants or when analyzing all participants together (Table S1). Comorbidities and receiving the BNT162b2 vaccine instead of the mRNA-1273 vaccine were associated with lower IgG levels (Table S1) and plasma neutralizing capacity (Table S2). Having been previously exposed, having had systemic AEs compared to local AEs or no AEs after the 1^st^ dose (for all and pre-exposed HCW) or the 2^nd^ dose (for all and naïve HCW) and days since the 1^st^ dose, were associated with higher IgG levels (Table S1). Curiously, IgG levels against the N protein of the HCoV HKU were negatively associated with post vaccination IgG levels against S full length in pre-exposed participants (Table S1). Being a smoker was also associated with lower plasma neutralizing capacity, while systemic AEs after the 2^nd^ dose were associated with higher neutralizing capacity (Table S2).

In stepwise multivariable models, age and sex were not significantly associated with IgG levels against S protein (Table 2). Previous SARS-CoV-2 exposure was associated with 38% (13-69%, 95% CI) higher IgG levels to S, whereas BNT162b2 vaccination was associated with 43% (31-54%, 95% CI) less IgG-S levels than mRNA-1273 vaccination, regardless of exposure. In addition, in all participants and in the unexposed ones, having had systemic AEs compared to local or no AEs after the 2^nd^ dose was associated with 23-28% higher IgG-S levels. In the pre-exposed HCW, being a smoker or having underlying comorbidities were independently associated with 35% (3-57%, 95% CI) and 55% (33-70%, 95% CI) less IgG-S levels, whereas there was a trend towards higher IgG-S levels when the HCW had a symptomatic infection compared to an asymptomatic infection.

Being smoker, having comorbidities, and receiving the BNT162b2 vaccine instead of the mRNA-1273 vaccine were also associated with 43%, 45% and 30% lower plasma neutralizing capacity, respectively (Table 3). Having had systemic AEs compared to local or no AEs after the 2^nd^ dose was associated with 60.54% higher neutralizing capacity. SARS-CoV-2 exposure was also associated with 30% higher plasma neutralizing capacity though the statistical significance was borderline (P-value=0.051).

**Table 3.** Step-wise multivariable models assessing the impact of several variables on the plasma neutralizing capacity after two doses of mRNA vaccines (12-19 days post-vaccine).

|  | Beta (%)<sup>a</sup> | Lower<br>95% CI | Upper<br>95% CI | P-value |
| --- | --- | --- | --- | --- |
| **Neutralizing capacity** |  |  |  |  |
| **All participants N=92** |  |  |  |  |
| Smoker | -42.80 | -59.47 | -19.27 | **0.002** |
| Comorbidities | -44.81 | -62.76 | -18.22 | **0.003** |
| SARS-CoV-2 exposure | 30.13 | -0.06 | 69.44 | 0.051 |
| BNT162b2 (ref: mRNA-1273) | -30.19 | -47.80 | -6.64 | **0.016** |
| Systemic AEs dose 1 (ref: local/no AEs) | 24.85 | -6.21 | 66.18 | 0.127 |
| Systemic AEs dose 2 (ref: local/no AEs) | 60.54 | 16.62 | 121.00 | **0.004** |

<sup>a</sup> Beta transformed values to a percentage for an easier interpretation of variables effect

### IgG levels to S antigens induced by natural infection are maintained for up to a year

Antibody kinetics since the onset of symptoms along 6 time-points for 102 exposed non-vaccinated individuals are shown in Fig. 6. At study month 12, 53 of the 414 HCW who visited had not received any vaccine dose yet, 36 of whom had been previously infected by SARS-CoV-2. IgM levels rapidly fell below the seropositivity thresholds. Similarly, IgA against full length N and its C-term region and IgG against N C-term decayed over time below the seropositivity thresholds. On the contrary, IgG and IgA levels against any of the S antigens tested (RBD, S or S2) remained positive over time for most of the participants for up to 1 year of follow-up, with IgG at higher levels than IgA. There were 31 exposed individuals with more than 300 days post-infection who had not been vaccinated. IgM, IgA and IgG seropositivity was 12.9%, 64.5% and 90.3%, respectively, for any of the antigens tested.

**Figure 6.**
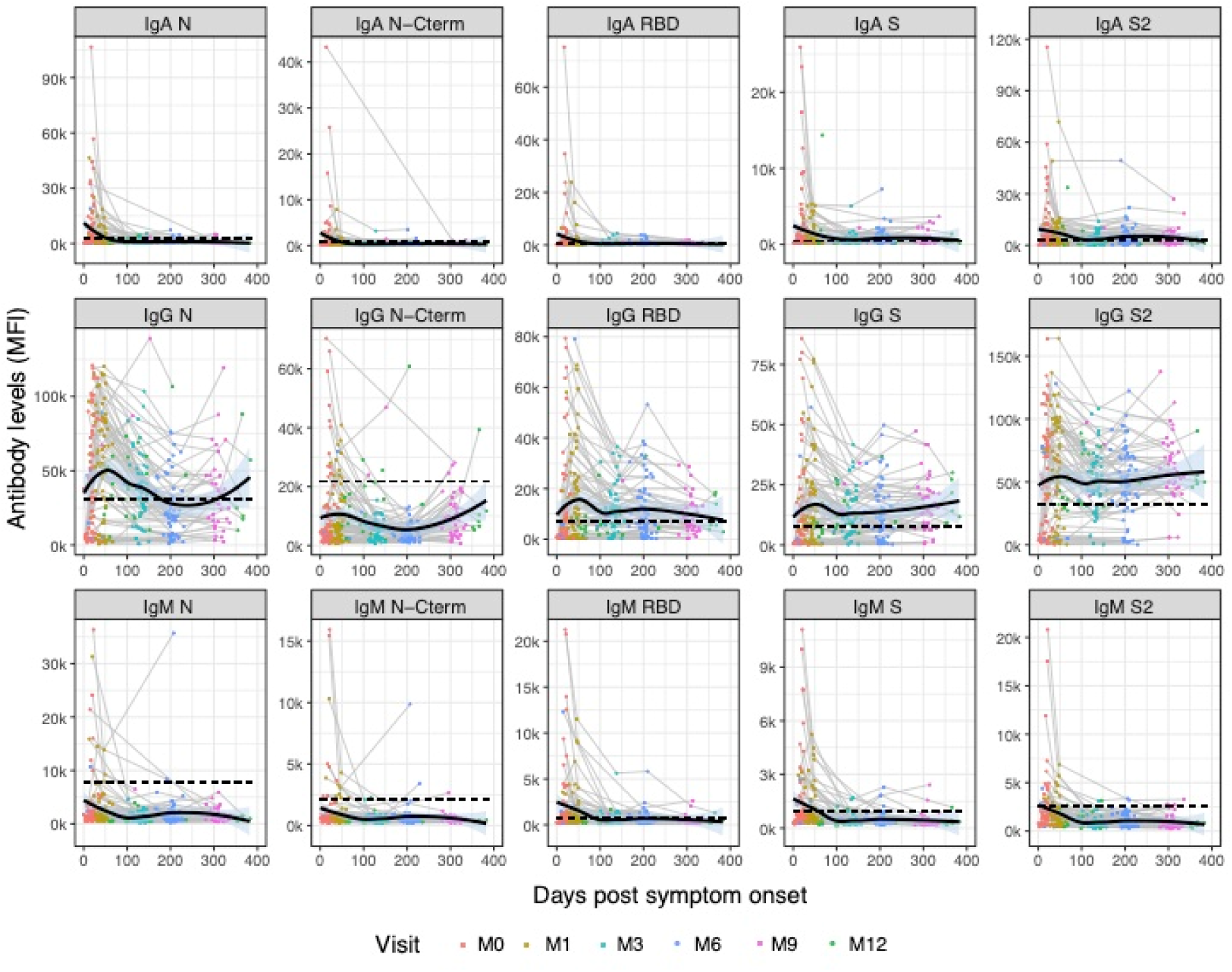
Kinetics of SARS-CoV-2 antibody levels since onset of symptoms in non-vaccinated participants. Levels (median fluorescence intensity, MFI) of IgA, IgG and IgM against each antigen (Nucleocapsid full length protein (N), and its C-terminal domain, the Receptor Binding Domain (RBD), full S protein and its subunit S2) measured in 338 samples from 102 symptomatic participants collected in up to 6 time points per participant (paired samples joined by lines). The black solid line represents the fitted curve calculated using the LOESS (locally estimated scatterplot smoothing) method. Shaded areas represent 95% confidence intervals. Dashed line represents the positivity threshold. Samples were analyzed at the 1:500 dilution.

## DISCUSSION

Knowledge on the antibody response induced by COVID-19 vaccines and the factors affecting it, such as previous SARS-CoV-2 infection, is essential to understanding immunity elicited by vaccination and its heterogeneity in the general population, which can be used to improve the design of vaccination policies and guide personalized recommendations. We analyzed IgA, IgG and IgM responses to the COVID-19 mRNA vaccines mRNA-1273 and BNT162b2 in a well-characterized cohort of HCW with detailed demographic and clinical information, accurate history of SARS-CoV-2 exposure, and antibody responses since the beginning of the pandemic. Our results show that COVID-19 mRNA vaccines induce robust antibody responses to S antigens in most of the HCW but mRNA-1273 elicited higher antibody levels and quality than BNT162b2. Independently of the vaccine received, antibody responses were higher in previously SARS-CoV-2 exposed individuals, particularly if they had a symptomatic infection, and a 2^nd^ dose of the vaccine in pre-exposed individuals did not increase their antibody levels, supporting the strategy of a single-dose vaccination for previously infected individuals to achieve a higher vaccination coverage and in more populations. However, our data also highlights the need for more personalized strategies as antibody responses may be diminished in asymptomatic, smokers and individuals with chronic diseases.

Higher IgG responses induced by mRNA-1273 than BNT162b2 have also recently been reported by others [47, 48], but to our knowledge, we are the first to report higher neutralizing capability and higher antibody avidity. The type of vaccine was not randomly administrated, but HCW were not allowed to choose the vaccine brand since this depended on vaccine availability and did not follow a pre-established pattern. In addition, results were adjusted by relevant confounders such as previous SARS-CoV-2 exposure. Albeit very high antibody levels are induced by both vaccines, differences may be relevant for those individuals responding more poorly to vaccination or naive individuals. Although both vaccines use the same technology, they differ in the amount of mRNA per dose (100 μg vs 30μg) [2, 3] and formulation, but also in the schedule of the 2^nd^ dose: 4 weeks after 1^st^ dose for mRNA-1273 vs. 3 weeks for BNT162b2, which could be related to the differences observed here. A delay in the 2^nd^ dose of the COVID-19 vaccine AstraZeneca (ChAdOx1-SARS-COV-2) showed improved immunogenicity and protection [49, 50]. This suggests that there may be room for optimization in the dose quantity and schedules.

Upon vaccination, exposed participants had higher levels of IgG and IgA against S antigens than naive participants after one dose of the vaccine and, after two vaccine doses, naive individuals still had lower IgG and IgA levels against S antigens and of lower neutralizing capacity and avidity than exposed individuals. As mentioned, the 2^nd^ dose seemed to not be beneficial in expanding the antibody response further, similarly to what has been reported by others [7,8,11]. Nevertheless, antibody responses were very heterogeneous, even among previously exposed individuals. We found that HCW who had an asymptomatic infection tended to ahve less IgG levels after vaccination than symptomatic HCW, and that SARS-CoV-2 antibodies before vaccination positively correlated with post-vaccination levels. Smokers and individuals with underlying comorbidities had considerably lower antibody levels and lower plasma neutralizing capacity. Therefore, a 2^nd^ dose should be considered for exposed individuals who were asymptomatic, are smokers or people with comorbidities, especially the immunosuppressed, or could be administered depending on previous antibody titers, although this approach depends on the identification of correlates of protection and would only be feasible in high-income countries.

Nevertheless, receiving the full schedule for exposed individuals may be relevant for maintenance of responses over time assuming a decline of antibodies [51], and to overcome the impact of new VoCs with increased transmissibility and potential immune escape, like the Delta variant. We have detected a 6.3% vaccine breakthrough among fully vaccinated participants in between 15 and 189 days post second dose, probably related to the fifth wave of the pandemic, mostly caused by the Delta variant. Here, we have not detected major differences in the IgG and IgA responses between any of the VoCs tested and the wild-type S, in contrast to other studies reporting diminished sensitivity of neutralizing antibodies against the Beta and Gamma variants [27,28,30–33].

AEs have been associated with previous SARS-CoV-2 exposure [22, 47]. Here we found that AEs, particularly after the 2^nd^ dose, were positively associated with antibody levels and neutralizing capacity, independently of having had previous SARS-CoV-2 exposure. Another study found that clinically significant reactions to these mRNA vaccines were associated to higher IgG levels [47]. AEs may reflect a strong innate response resulting in increased acquired responses.

Despite the clear impact of SARS-CoV-2 exposure on vaccines responses, time since infection did not have a major effect. In face of shortage of vaccine doses, and based on some studies reporting maintenance of antibody responses in COVID-19 recovered patients for more than 6 months [52–55] recommendations to wait up to 6-month post-infection to get vaccinated were issued in some countries, including Spain [23]. Nevertheless, at HCB, all HCW were recommended to get the vaccine although naïve individuals were prioritized.

Here, we show maintenance of IgG responses up to a year post-infection. After more than 300 days (up to 383 days) following infection among unvaccinated HCW, 90% were still seropositive for IgG against any of the S antigens, demonstrating persistence of immunity to natural exposure. This suggests maintenance of a certain level of protection irrespective of the additional role of memory T cell responses [52, 54]. Based on our data, it is difficult to recommend how long previously exposed individuals could wait to get vaccinated, although receiving at least a dose of a mRNA vaccine if previously exposed clearly increases antibody levels and neutralizing capacity regardless of the time since infection.

Currently approved mRNA COVID-19 vaccines have proven to be highly efficacious [2, 3] and effective in the real population [12–15] for at least a few months but durability of immunity and vaccine escape by VoCs needs to be monitored. Optimal antibody responses elicited early after vaccination may be important for maintenance of immunity and protection and we have demonstrated that responses depend on the vaccine received, number of doses, previous history of SARS-CoV-2 infection and SARS-CoV-2 immune responses, life style and health of the individuals. Even in a cohort of HCW, we have found a high heterogeneity of antibody responses and highlight the need of more personalized recommendations. Moving forward, differential quantitative and qualitative responses to the vaccines between exposed and naive individuals from different populations and conditions needs to be studied over time to better inform vaccination strategies.

## Data Availability

The antibody levels, avidity and neutralization data generated in this study and metadata will be deposited in the UB repository upon publication of the manuscript.

## Author contributions

G.M., R.A., A.L.G-B., C.D. designed the study. G.M., R.A. and C.D. supervised all work. R.A., C.C., P.V., S.B., M.T. and C.D. coordinated participant visits and sample and data collection. G.S., R.A., D.B, M.V., C.D. and L.P. collected samples and data at HCB. R.R., MJ.M., M.V., D.B., R.A.M. and A.J. processed the samples, developed and performed the serological Luminex assays and analyses. P.H-L, A.A. and P.E. designed and performed the flow cytometry neutralization assay. N.R.M., C.R. and L.I. produced the antigens. A.LL., A.M., A.T., A.V. contributed to design and the critical interpretation of the results. N.O., M.R., and P.R. managed and analyzed the data and N.O., M.R. and G.M. prepared the manuscript figures. S.M. managed the clinical data. G.M., R.A., N.O., M.R., C.D. interpreted the results and wrote the first draft of the paper. G.M. R.A. N.O, M.R., C.D. had access to, and verified, the data. G.M. and R.A. contributed equally. M.R., N.O., R.R., G.S. also contributed equally. A.L.G-B. & C.D. jointly supervised this work. All authors approved the final version as submitted to the journal.

## Acknowledgments

We thank the participation of HCW who are committed to this study and are key personnel facing the pandemic. We are grateful to Pau Cisteró, Chenjerai Jairoce, Selena Alonso, Rebeca Santano, Sarah Williams, Montserrat Lamoglia, Neus Rosell, Angeline Cruz, Eugénia Chóliz, Antía Figueroa-Romero, Silvia Folchs, Jochen Hecht, Mikel J. Martínez, Núria Pey, Patricia Sotomayor and Sara Torres who participated in the field and/or laboratory work during previous visits. We also thank the administrative department in ISGlobal, and Gemma Ruiz-Olalla, and Sergi Sanz for statistical advice. Maria José Molina was registered in the EMJMD LIVE (Erasmus+ Mundus Joint Master Degree Leading International Vaccinology Education), cofunded by the EACEA (Education, Audiovisual and Culture Executive Agency, award 2015-2323) of the European commission, and received a scholarship from the EACEA.

## Data Sharing Statement

The antibody levels, avidity and neutralization data generated in this study and metadata are deposited in the UB repository under this link: XXXXX.

## Supplementary Material

### Supplementary Methods

#### Quantification of antibodies to SARS-CoV-2 by Luminex

The levels of IgG, IgM and IgA were assessed in single replicates by high-throughput multiplex quantitative suspension array technology (qSAT). The assay was performed in 5 plates of 384 wells for IgG, and 3 plates for IgA and IgM with samples from the same individual in the same plate. Assay performance was previously established as 100% specificity and 95.78% sensitivity for seropositivity 14 days after symptoms onset [1].

#### Coupling of proteins to microspheres

MagPlex® polystyrene 6.5 μ COOH-microspheres (Luminex Corp, Austin, TX, USA) were washed, sonicated and activated with Sulfo-NHS (N-hydroxysulfosuccinimide) and EDC (1-Ethyl-3-[3-dimethylaminopropyl]carbodiimide hydrochloride) (Thermo Fisher Scientific Inc., Waltham USA). Next, microspheres were washed and resuspended in 50 mM MES pH 5.0 (MilliporeSigma, St. Louis, USA). The recombinant proteins were then incubated with the microspheres at the optimal concentrations (from 10 to 50 μ mL) and left at room temperature on a shaker for two hours. Coupled microspheres were resuspended in PBS with 1% BSA and 0.05% Tween 20 to covalently block the free carboxylic group (-COOH) absorbing most of the non-specific binding to secondary antibodies during assay steps [2] and heterophilic antibody binding seen in previous systems [3]. Microspheres recovery was quantified on a Guava® easyCyte™ Flow Cytometer (Luminex Corporation, Austin, USA). Equal amounts of each antigen-coupled microspheres were multiplexed and stored at 2000 microspheres/μL at 4°C, protected from light.

#### qSAT assay

Antigen-coupled microspheres were added to a 384-well Clear® flat bottom plate (Greiner Bio-One, Frickenhausen, Germany) in multiplex (2000 microspheres per analyte per well) in a volume of 90 μL of Luminex Buffer (1% BSA, 0.05% Tween 20, 0.05% sodium azide in PBS) using 384 channels Integra Viaflo semi-automatic device (96/384, 384 channel pipette). Two hyperimmune pools (one for IgG, and another one for IgA and IgM) were used as positive controls in each assay plate for QA/QC purposes and were prepared at 2-fold, 8 serial dilutions from 1:12.5. Pre-pandemic samples were used as negative controls to estimate the cut off of seropositivity. Ten μL of each dilution of the positive control, negative controls and test samples (prediluted 1:50 in 96 round-bottom well plates), were added to a 384-well plate using Assist Plus Integra device with 12 channels Voyager pipette (final test sample dilution of 1:500). To quantify IgM and IgA responses, test samples and controls were pre-treated with anti-Human IgG (Gullsorb) at 1:10 dilution, to avoid IgG interferences. Technical blanks consisting of Luminex Buffer (PBS with 1% BSA) and microspheres without samples were added in 4 wells to detect and adjust for non-specific microsphere signals. Plates were incubated for 1 h at room temperature in agitation (Titramax 1000) at 900 rpm and protected from light. Then, the plates were washed three times with 200 μ /well of PBS-T (0.05% Tween 20 in PBS), using BioTek 405 TS (384-well format). Twenty five μ of goat anti-human IgG phycoerythrin (PE) (GTIG-001, Moss Bio) diluted 1:400, goat anti-human IgA-PE (GTIA-001, Moss Bio) 1:200, or goat anti-human IgM-PE (GTIM-001, Moss Bio) 1:200 in Luminex buffer were added to each well and incubated for 30 min. Plates were washed and microspheres resuspended with 80 μ of Luminex Buffer, covered with an adhesive film and sonicated 20 seconds on sonicator bath platform, before acquisition on the Flexmap 3D® reader. At least 50 microspheres per analyte per well were acquired, and median fluorescence intensity (MFI) was reported for each analyte.

## Supplementary figures

**Figure S1.**
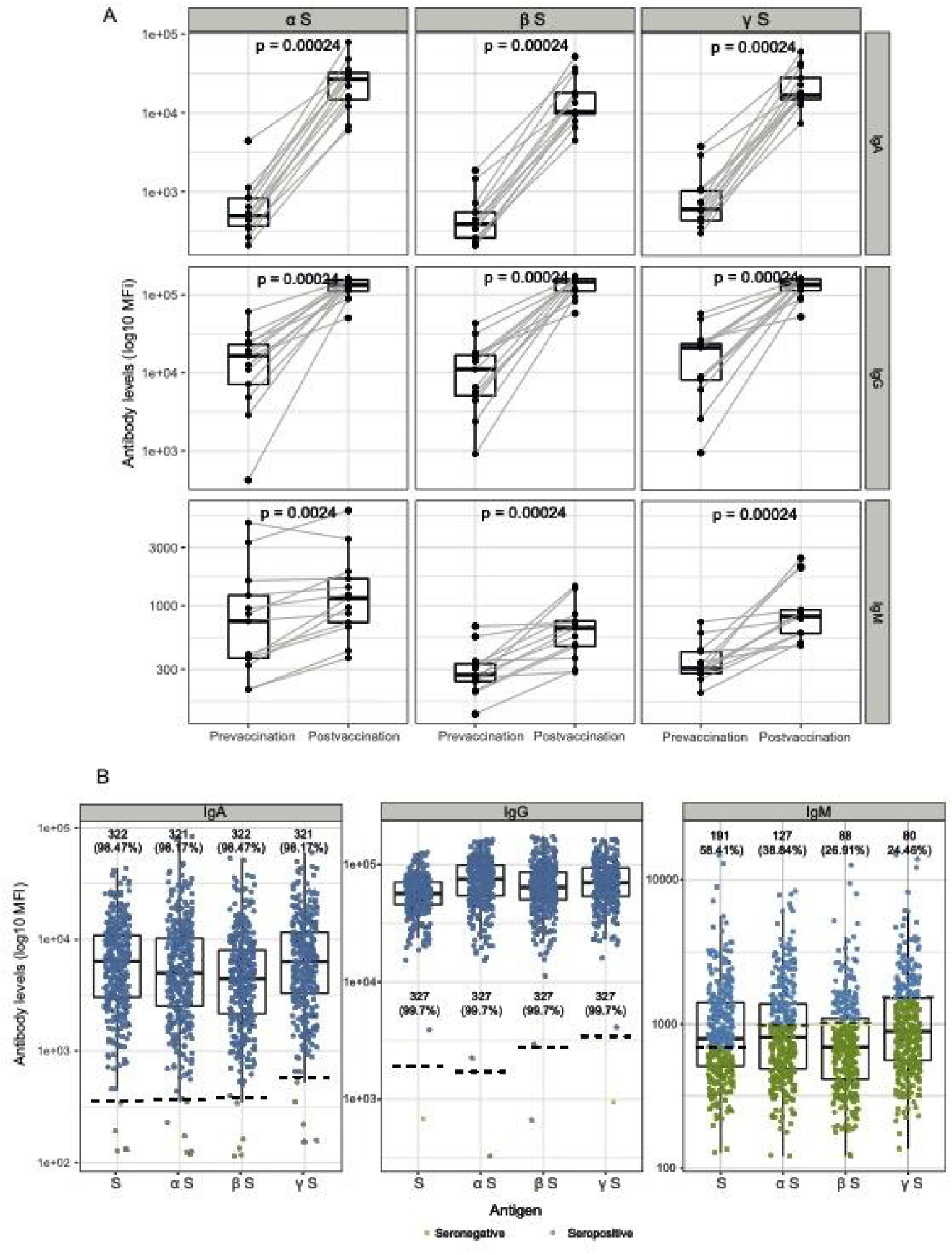
Antibody levels and seropositivity against the S antigen from the Wuhan and the Alpha, Beta and Gamma variants of concern. The graphs show IgA, IgG and IgM levels (median fluorescence intensity, MFI) against the Spike protein of Alpha, Beta and Gamma variants A) before and after two vaccine doses in a subset of individuals with available pre-vaccination data (N=13, 12 mRNA-1273 and 1 BNT162b2) at M9 (1:500 dilution for pre and 1:5000 for post) and B) after two vaccine doses in all participants (N=328). The center line of boxes depicts the median of MFIs; the lower and upper hinges correspond to the first and third quartiles; the distance between the first and third quartiles corresponds to the interquartile range (IQR); whiskers extend from the hinge to the highest or lowest value within 1.5 × IQR of the respective hinge. In A, paired samples are joined by grey lines and Wilcoxon signed-rank test was used to assess statistically significant differences in antibody levels between pre- and post-vaccination. In B, dashed lines represent the positivity threshold, and numbers and percentages indicate the number and proportion of seropositive participants for each isotype and S protein.

**Figure S2.**
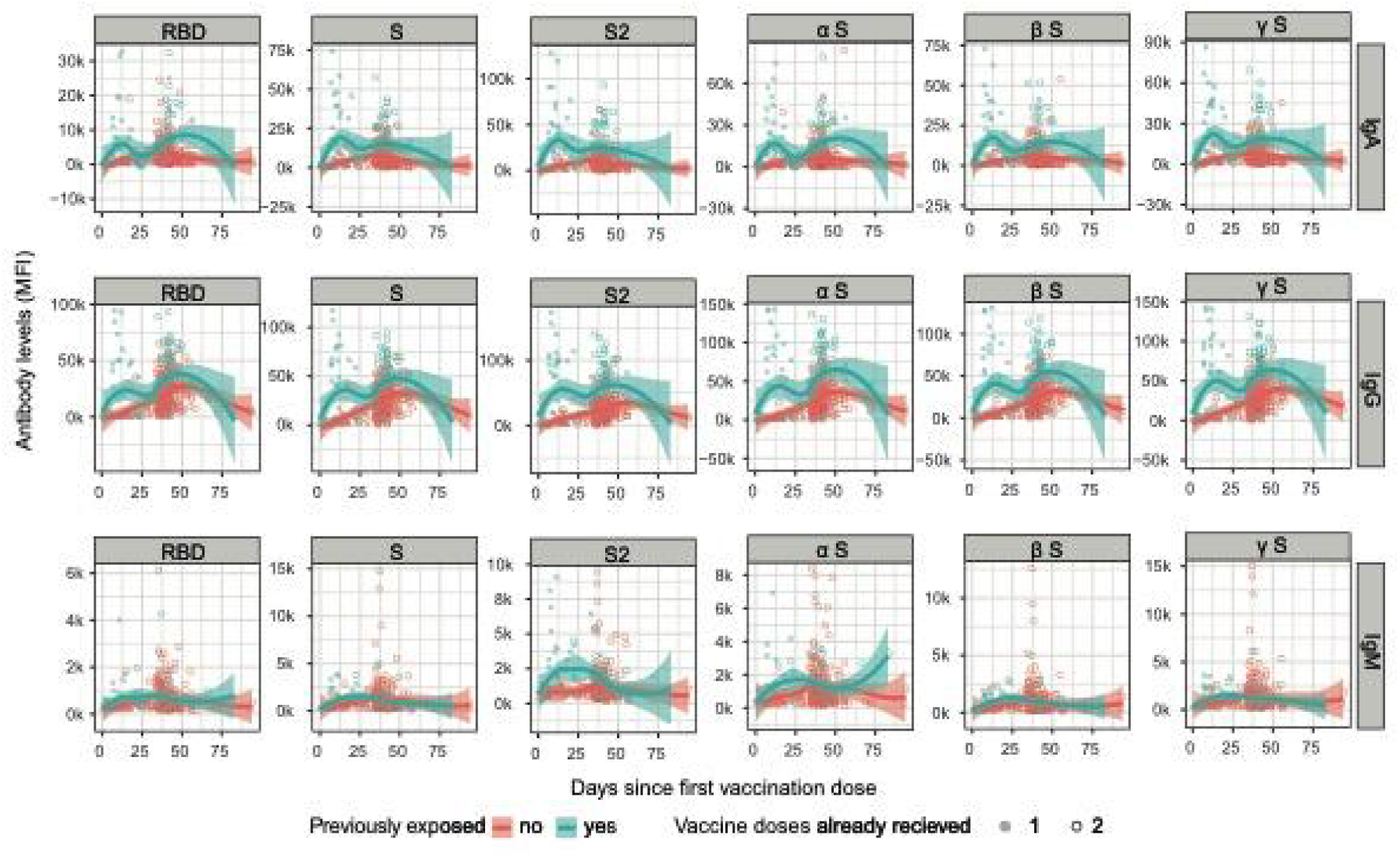
Kinetics of antibody levels against the S antigens since 1^st^ vaccination dose in naive and pre-exposed participants after 1^st^ and 2^nd^ doses. Levels (median fluorescence intensity, MFI) of IgA, IgG and IgM against each antigen (the Receptor Binding Domain (RBD), full S protein, its subregion S2 and full S protein of the variants of concern) measured in 426 samples from 368 participants. The red and green solid lines represent the fitted curve calculated using the LOESS (locally estimated scatterplot smoothing) method. Shaded areas represent 95% confidence intervals. Red and Green shaded areas correspond to naive and pre-exposed participants, respectively. Filled and empty points correspond to samples collected after 1 and 2 doses, respectively. Samples were analyzed at the 1:5000 dilution.

**Figure S3.**
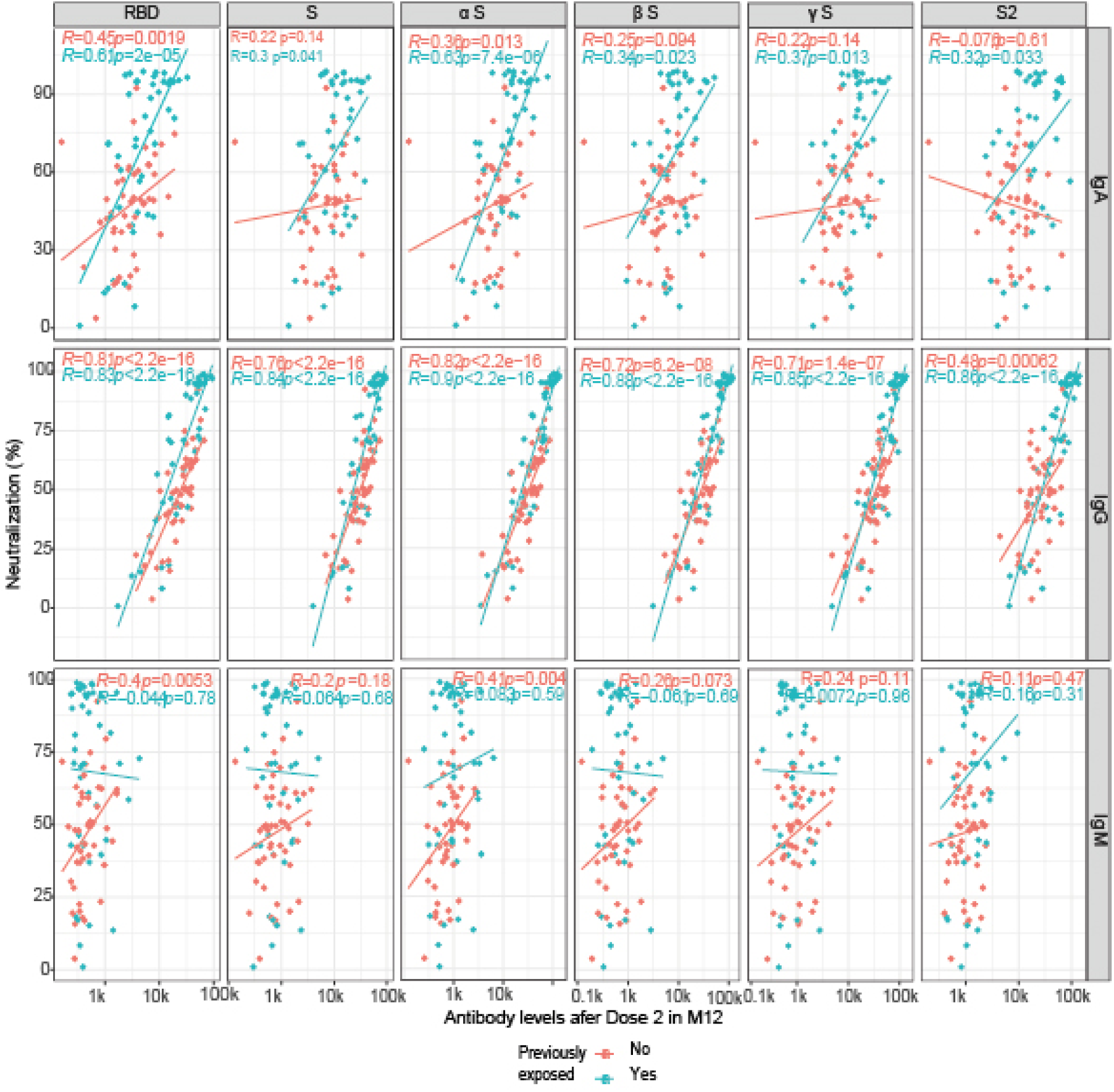
Correlations between plasma neutralization activity and antibody levels after 2 vaccine doses in naive and exposed participants. Scatter plots showing the correlation between plasma neutralization capacity (as a percentage of RBD-ACE2 binding inhibition) and IgA, IgG and IgM levels (median fluorescence intensity, MFI) against each antigen (the Receptor Binding Domain (RBD), full S protein, its subregion S2 and full S proteins of the variants of concern). P-values and rho correlation coefficients were computed through Spearman’s rank correlation tests. Lines represent the fitted curve calculated using the linear model method. Red and green dots and lines correspond to naive (N=47) and pre-exposed (N= 45) participants, respectively. A dilution of 1:400 was used for the neutralization assay, 1:500 for IgA and IgM levels, and 1:5000 for IgG levels.

**Figure S4.**
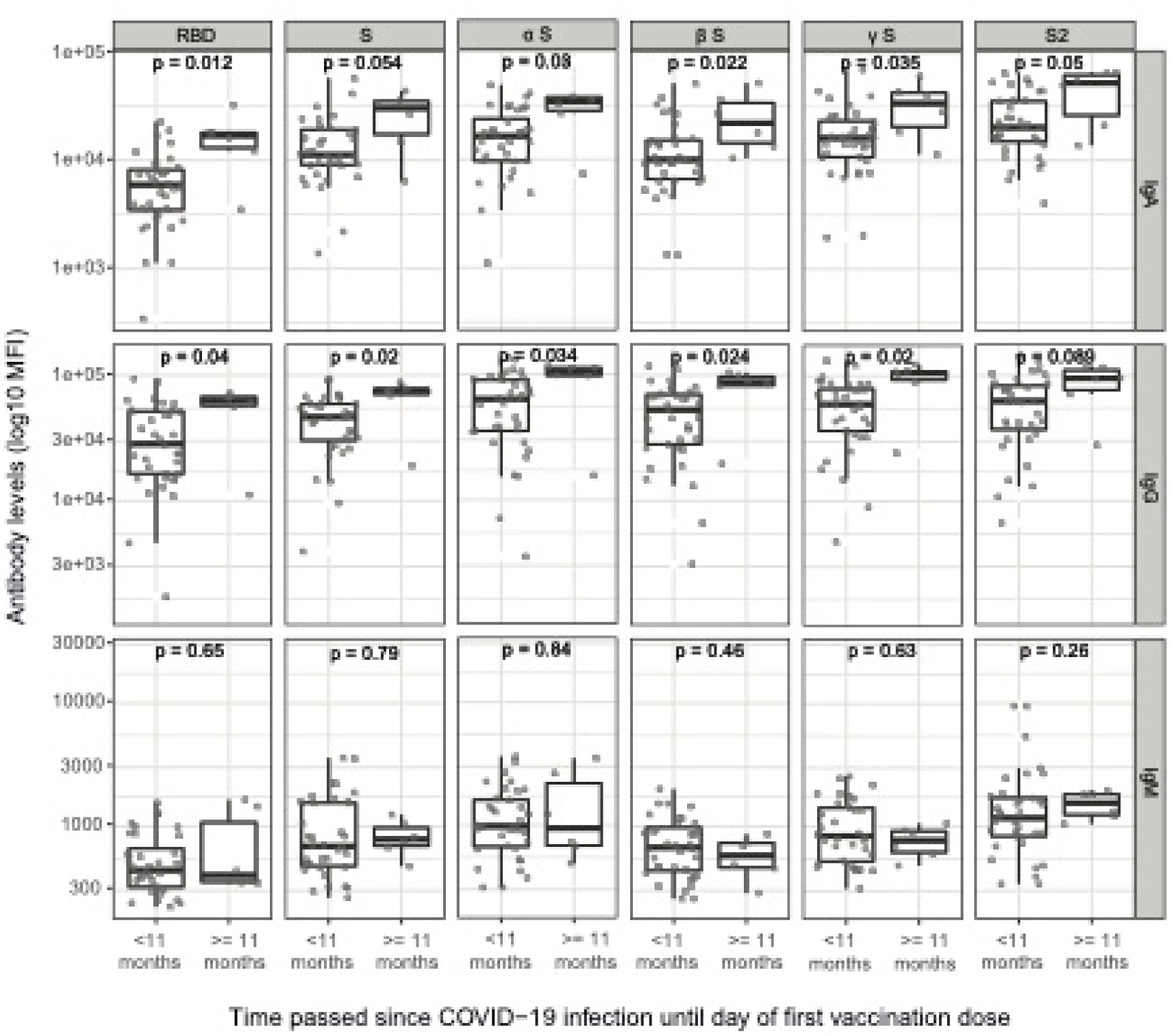
Comparison of antibody levels to S antigens after two COVID-19 mRNA vaccine doses between participants infected more vs less than 11 months prior to vaccination. Plots show IgA, IgG and IgM levels (log_10_MFI) against the receptor-binding domain (RBD) of the SARS-CoV-2 Spike glycoprotein (S), the full S protein, its subunit S2 and full S proteins for the variants of concern in participants infected more (N=6) or less (N=34) than 11 months prior to vaccination. The center line of boxes depicts the median of MFIs; the lower and upper hinges correspond to the first and third quartiles; the distance between the first and third quartiles corresponds to the interquartile range (IQR); whiskers extend from the hinge to the highest or lowest value within 1.5 × IQR of the respective hinge. Wilcoxon rank test was used to assess statistically significant differences in antibody levels between groups.

**Figure S5.**
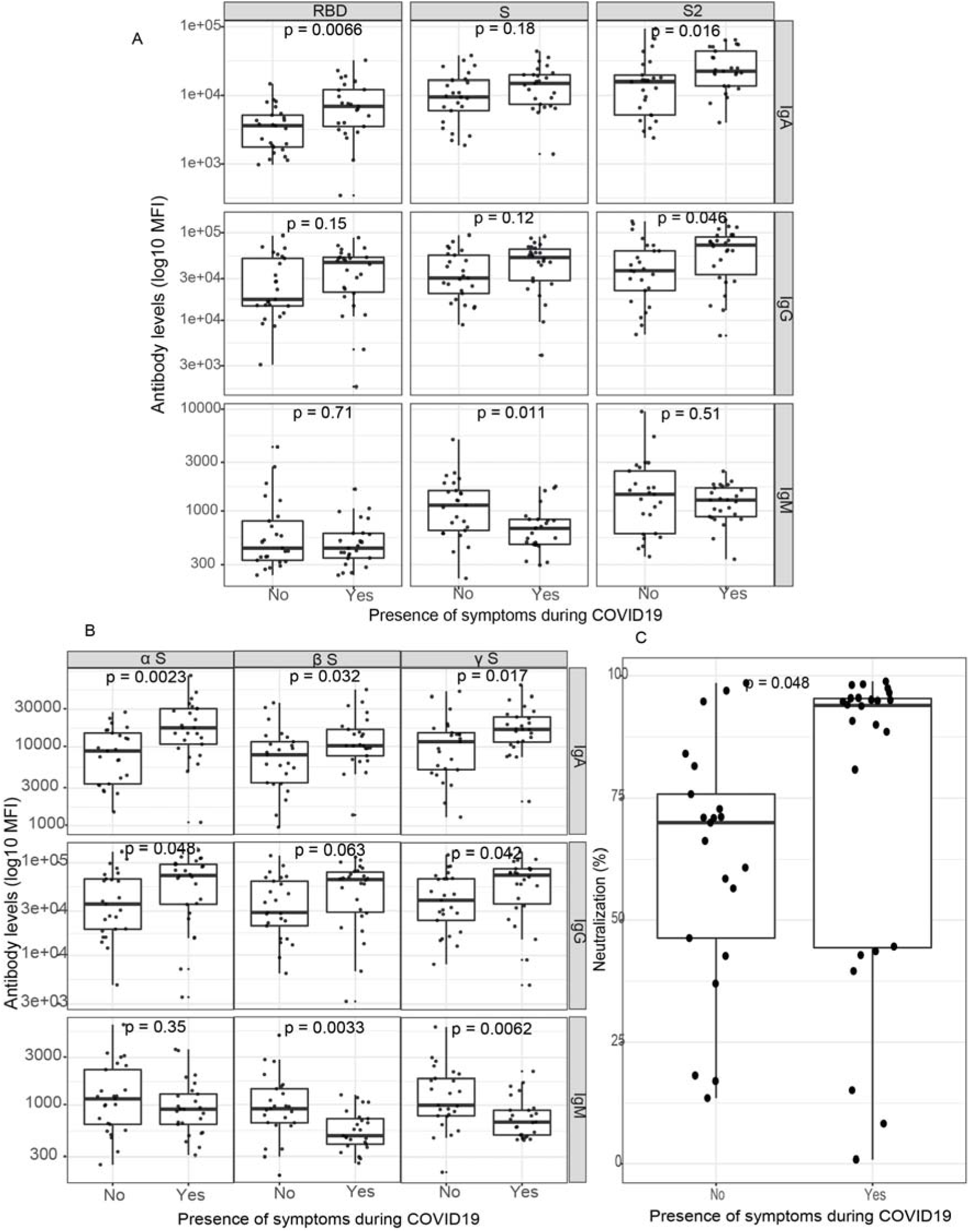
Comparison of antibody levels to S antigens and plasma neutralization capacity after two COVID-19 mRNA vaccine doses between symptomatic and asymptomatic pre-exposed participants. Antibody levels to spike (S) antigens from Wuhan strain (A) and to S proteins from the variants of concern (B) (total N= 51, 25 asymptomatic and 26 symptomatic). Plasma dilutions tested were 1:500 for IgA/M and 1:5000 for IgG. C) Plasma neutralizing capacity after two vaccine doses (N=45) in previously infected symptomatic (N=25) and asymptomatic (N=20) participants (dilution tested was 1:400).

**Figure S6.**
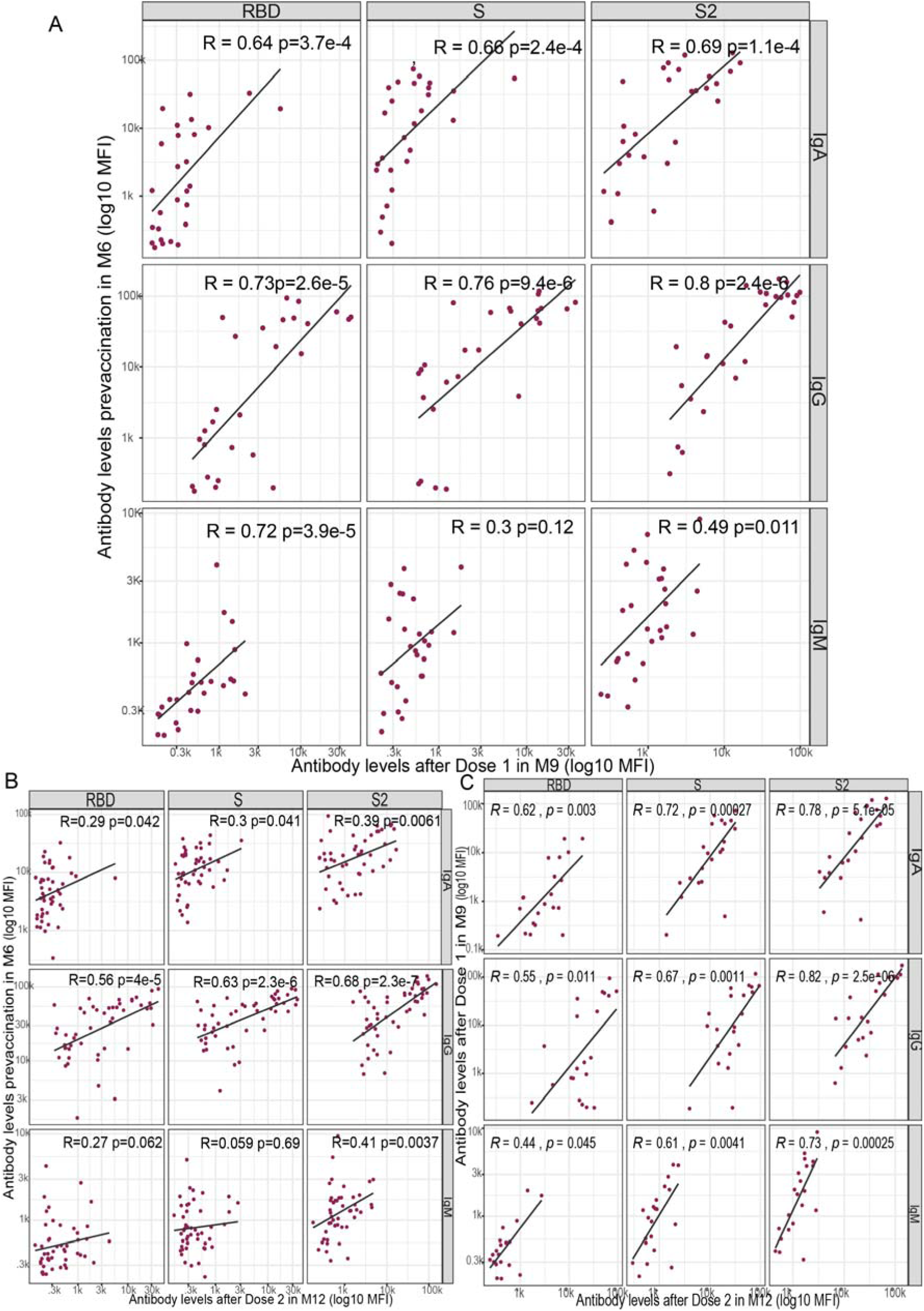
Correlation of antibody levels before and after COVID-19 mRNA vaccination and between doses in pre-exposed participants. A) Correlations between pre-vaccination levels at M6 and >7 days post dose 1 (BNT162b2, N=27). Sample dilution analyzed was 1:500. B) Correlations between pre-vaccination (M6) and post-vaccination antibody levels after 2 doses (BNT162b2 and mRNA-1273 pooled, N=49). Sample dilution analyzed was 1:500 for IgA/M and 1:5000 for IgG. C) Correlations between antibody levels post dose 1 (1:500 dilution) and post dose 2 (1:500 dilution for IgA/M and 1:5000 for IgG); samples post dose 1 used in the analysis were collected >7 days post vaccination (N=21).

**Figure S7.**
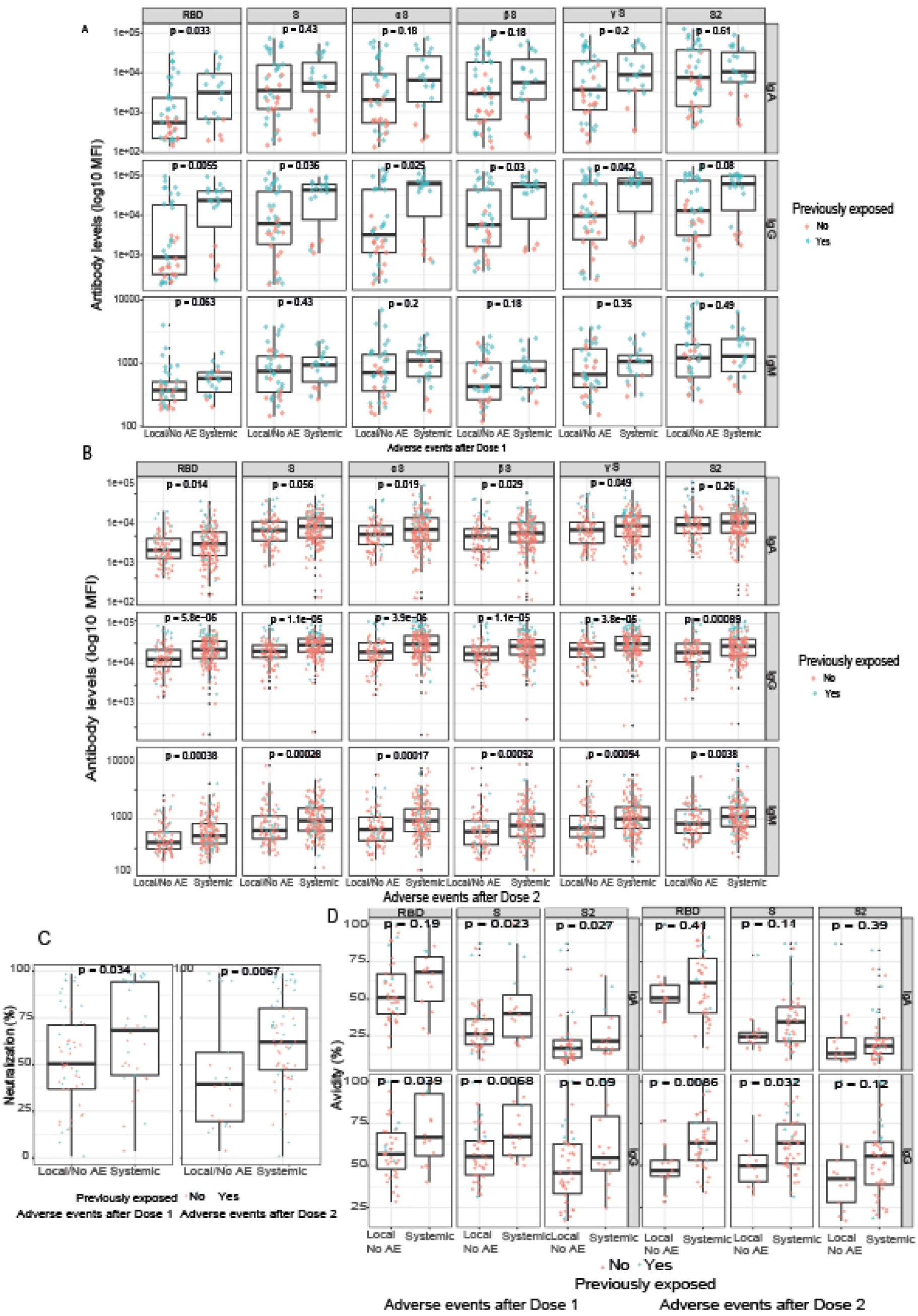
Comparison of antibody levels and avidity to S antigens and plasma neutralization capacity between participants with systemic vs local or no adverse events after vaccination. Antibody levels after 1^st^ dose (N=57, Systemic N=19, Local/No AEs N=38, plasma 1:500 dilution) (A) and 2^nd^ dose (N=263, Systemic N=84, Local/No AE N =179, plasma 1:500 dilution for IgA/IgM and 1:5000 for IgG) (B). Plasma neutralization capacity (N=92, 1:400 dilution) after dose 1 (Systemic N=36, Local/No AE N=56) and at dose 2 (Systemic N=67, Local/No AE N=25) (C), and avidity (N=58, 1:5000 dilution) after dose 1 (Systemic N=16, Local/No AE N= 42) and dose 2 (Systemic N= 45, Local/No AE N=13) (D). Red and green dots correspond to naive and pre-exposed participants, respectively.

**Table S1.**
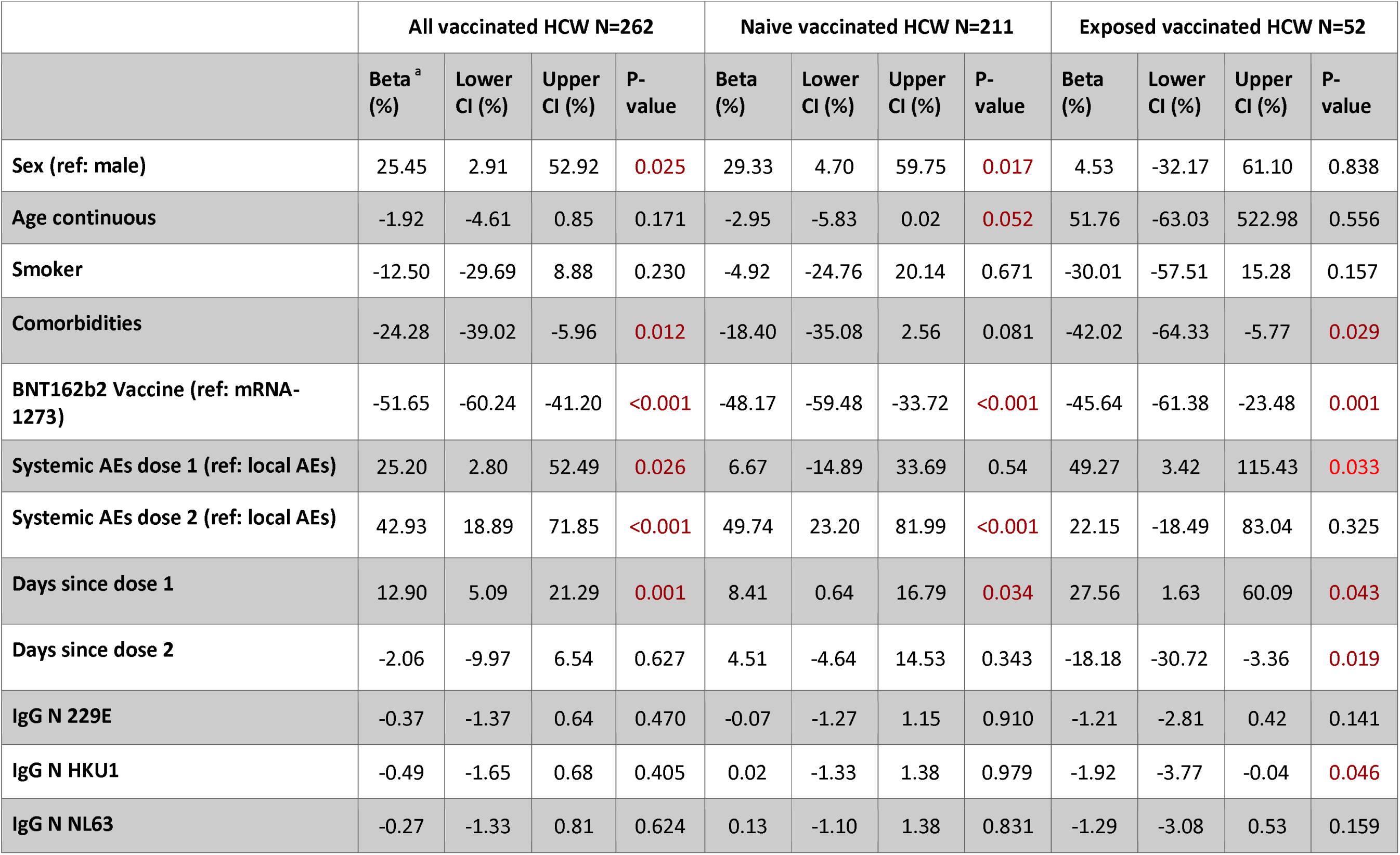
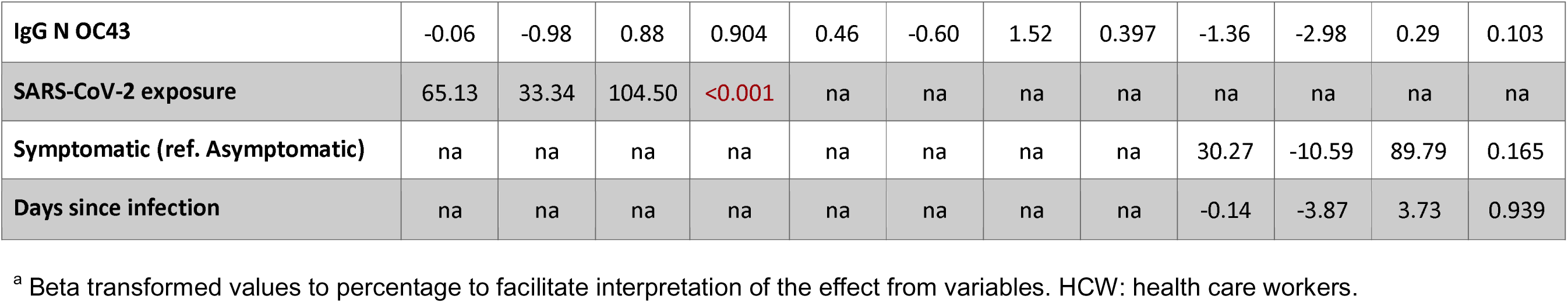
Univariable models assessing variables affecting IgG S levels after two doses of the mRNA vaccines.

**Table S2.** Univariable models assessing variables affecting the plasma neutralization capacity (%) after two doses of the mRNA vaccines in all vaccinated health care workers (N=92).

|  | All vaccinated HCW N=92 |  |  |  |
| --- | --- | --- | --- | --- |
|  | Beta (%)<sup>a</sup> | Lower CI (%) | Upper CI (%) | P-value |
| Sex (ref: male) | 23.62 | -12.60 | 74.85 | 0.227 |
| Age continuous | -0.87 | -5.51 | 4.00 | 0.720 |
| Smoker | -37.81 | -57.81 | -8.32 | **0.017** |
| Comorbidities | -38.24 | -60.64 | -3.09 | **0.036** |
| BNT162b2 Vaccine (ref: mRNA-1273) | -40.71 | -55.69 | -20.68 | **0.001** |
| Systemic AEs dose 1 (ref: local AEs) | 29.58 | -5.33 | 77.37 | 0.104 |
| Systemic AEs dose 2 (ref: local AEs) | 55.13 | 10.72 | 117.34 | **0.011** |
| Days since dose 1 | 9.94 | -1.65 | 22.90 | 0.095 |
| Days since dose 2 | -8.36 | -21.64 | 7.17 | 0.271 |
| IgG N 229E | -1.11 | -2.74 | 0.54 | 0.182 |
| IgG N HKU1 | -0.82 | -2.91 | 1.30 | 0.440 |
| IgG N NL63 | -1.02 | -2.80 | 0.80 | 0.266 |
| IgG N OC43 | -0.75 | -2.57 | 1.11 | 0.422 |
| **SARS-CoV-2 exposure** | 32.59 | -2.30 | 79.95 | 0.070 |

<sup>a</sup> Beta transformed values to % for an easier interpretation of the effect from variables.

